# Hyper-diverse antigenic variation and resilience to transmission-reducing intervention in falciparum malaria

**DOI:** 10.1101/2024.02.01.24301818

**Authors:** Qi Zhan, Qixin He, Kathryn E. Tiedje, Karen P. Day, Mercedes Pascual

## Abstract

Intervention against falciparum malaria in high transmission regions remains challenging, with relaxation of control efforts typically followed by rapid resurgence. Resilience to intervention co-occurs with incomplete immunity, whereby children eventually become protected from severe disease but not infection and a large transmission reservoir results from high asymptomatic prevalence across all ages. Incomplete immunity relates to the vast antigenic variation of the parasite, with the major surface antigen of the blood stage of infection encoded by the multigene family known as *var*. Recent deep sampling of *var* sequences from individual isolates in northern Ghana showed that parasite population structure exhibited persistent features of high-transmission regions despite the considerable decrease in prevalence during transient intervention with indoor residual spraying (IRS). We ask whether despite such apparent limited impact, the transmission system had been brought close to a transition in both prevalence and resurgence ability. With a stochastic agent-based model, we investigate the existence of such a transition to pre-elimination with intervention intensity, and of molecular indicators informative of its approach. We show that resurgence ability decreases sharply and nonlinearly across a narrow region of intervention intensities in model simulations, and identify informative molecular indicators based on *var* gene sequences. Their application to the survey data indicates that the transmission system in northern Ghana was brought close to transition by IRS. These results suggest that sustaining and intensifying intervention would have pushed malaria dynamics to a slow-rebound regime with an increased probability of local parasite extinction.

**One Sentence Summary:** Population genomics of hyper-diverse *var* genes inform resurgence dynamics in falciparum malaria.

## INTRODUCTION

High-transmission endemic regions present an important challenge to controlling and eliminating falciparum malaria. Although malaria prevalence has declined considerably in many parts of the world in the last two decades, high-transmission regions, found primarily in sub-Saharan Africa, tend to be highly resilient to intervention efforts (*1*). Here, malaria incidence often rebounds to pre-intervention levels once control is relaxed, and sustained efforts are limited not only by the availability of resources but also by the spread of resistance to insecticides and drug treatments (*2, 3*). Children experience clinical episodes, eventually becoming immune to severe disease but not infection (*4*). This results in a large reservoir of asymptomatic infections in hosts of all ages, which contributes to sustained disease transmission (*5, 6*). A similar transmission reservoir is found in other vector-borne diseases in domestic and wildlife hosts that exhibit high prevalence of infection but incomplete immunity with no clinical symptoms (*7, 8, 9, 10*).

In these pathogens, a high level of asymptomatic infection at high transmission rates and associated incomplete immunity are enabled by high antigenic variation encoded via multigene families (*11*). The *var* multigene family in the malaria parasite *Plasmodium falciparum* provides an example; it encodes for the major variant surface antigen (VSA) during the blood stage of infection, the protein PfEMP1 (*Plasmodium falciparum* erythrocyte membrane protein 1) (*12, 13, 14, 15*). Anti-*PfEMP1* immunity has been found to be crucial to prevent severe disease manifestations and to clear infection (*16, 17, 18, 19*). Each parasite carries 50-60 *var* genes across its chromosomes. They are expressed largely sequentially, producing and exporting different variants of this protein to the surface of red blood cells (*12, 20*). Specific immune memory of a given VSA allows the host immune system to clear parasitized red blood cells, shortening infection and reducing the likelihood of parasite transmission. In contrast, when a strain expresses many VSAs a host has not previously encountered, long infectious periods ensue, enhancing its chance of transmission to another host (*19, 12*). Under high-transmission settings, local parasite populations exhibit a vast pool of *var* gene variants, documented to reach in the thousands to tens of thousands (*21, 22, 23, 24*) and generated mostly through mitotic and meiotic recombination but also mutation, as well as from migration of the human host and the mosquito vectors (*25, 26, 27, 28*). Previous studies have shown that negative frequency-dependent selection (NFDS) mediated by the acquisition of specific immunity by hosts contributes to the limited overlap of the *var* genes of both individual repertoires (individual parasite genomes) and isolates (sets of individual parasite genomes co-infecting individual hosts), in a pattern that is both non-random and non-neutral (*23, 29*). Given the large number of possible *var* gene combinations, hosts are not likely to have seen all or many of the VSAs encoded by the different circulating genomes, and they therefore remain susceptible to re-infection throughout their lifetime despite previous repeated exposure (*24, 30*). Multi-genomic infections are common resulting from either successive mosquito bites (i.e., superinfection) or a single mosquito bite (i.e., co-transmission) (*31, 32, 33, 34*).

A recent longitudinal field study in Bongo District, located in northern Ghana, implemented deep sampling and sequencing of *var* genes from individual isolates to monitor the response of the transmission system to a three-round transient intervention of indoor residual spraying (IRS) with insecticides (*21, 35*). Sequencing specifically concerns the DBLα tag, a small conserved ∼450bp region within *var* genes which encodes for the immunogenic Duffy-binding-like alpha domain of PfEMP1 (*21, 22, 23, 24, 36, 37, 38*). Bioinformatic analyses of a large database of exon 1 sequences of *var* genes showed a predominantly 1-to-1 DBLα-*var* relationship, such that each DBLα tag typically represents a unique *var* gene (*39*). We use DBLα and *var* interchangeably hereafter. This surveillance through molecular epidemiology revealed persistent estimates of high *var* diversity and low parasite strain overlap (low repertoire similarity), characteristic features of high-transmission regions, despite a considerable decrease in prevalence and parasite population size (*21, 35*). The transmission system rebounded rapidly after the IRS was discontinued, with this transient intervention appearing to have had only a limited impact (*21*). Was this indeed the case? We examine here whether deep sampling of sequences of *var* genes in a host population can help us evaluate the impact of intervention further, from which we specifically infer whether sustaining and intensifying control efforts would have taken the system through a pronounced transition and averted the rapid rebound of prevalence.

To address these questions, we first investigate the response of the epidemiological system to transmission-reducing interventions with an extended stochastic agent-based model (ABM) that incorporates strain diversity from a multigene family and the interplay between parasite evolution and malaria population dynamics (Materials and Methods) (*29*). We address the existence of a threshold behavior with intervention intensity, whereby the perturbed system would lose its capacity to rebound rapidly, and that of molecular indicators of such a transition based on deep sampling of *var* gene sequences. We then apply the lessons learned from the computational model to field data from northern Ghana. Results indicate that the structure of diversity revealed via molecular surveillance can inform intervention against malaria in high-transmission endemic settings. These findings should be relevant to other pathogens of humans, wildlife, and livestock with similar immune evasion strategies based on hyper-diverse antigenic variation (*7, 8, 9, 10*).

## RESULTS

We investigate how different intervention schemes impact the population genomic structure under different transmission regimes (Materials and Methods, see Fig. 1A-D for the numerical simulation designs and scenarios considered). We start with sustained interventions of a decade to then consider shorter, more realistic, transient ones (Materials and Methods, Fig. 1C). Numerical simulations of the ABM demonstrate three qualitatively different rebound dynamics in prevalence (Fig. 2A, figs. S1A) across the series of sustained IRS interventions (Fig. 1C-D). Systems under low- or mid-coverage vector control rebound rapidly in prevalence back to levels similar to those pre-intervention, in what we refer to as a fast-rebound regime. We use “rebound” hereafter synonymously with “resurgence” from the malaria literature, both describing a return to a baseline. In contrast, systems under high-coverage interventions remain at low prevalence, entering what we describe as a slow-rebound regime. Lastly, systems under interventions with levels between these two extremes gradually settle into intermediate prevalence, exhibiting a “transition” regime.

**Fig. 1.**
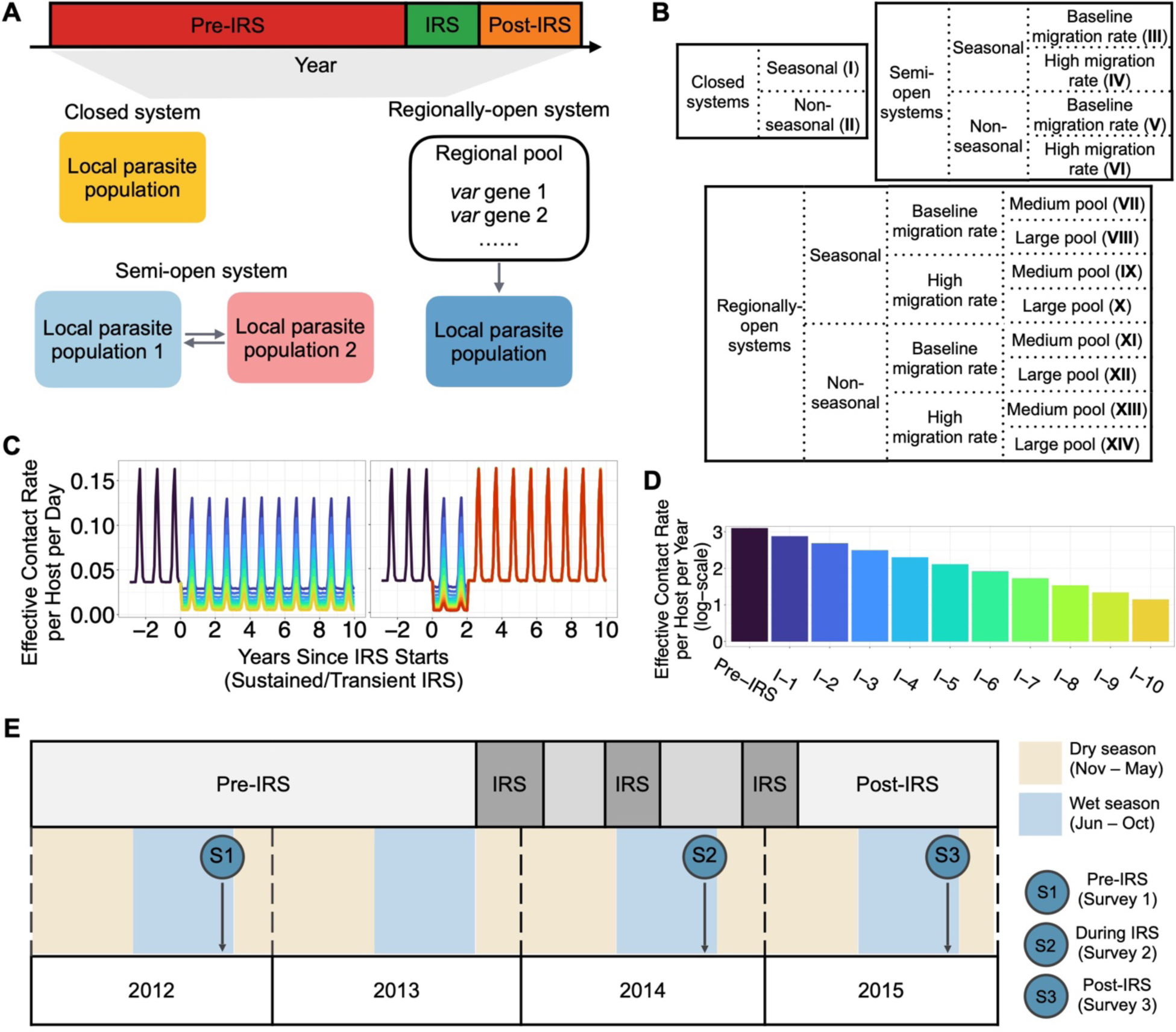
Diagram of the numerical simulation design mimicking different lengths of the transmission-reducing intervention (the application of indoor residual spraying or IRS) and the study design of field data sampled from Bongo District in northern Ghana. (**A**) Each simulation follows either two or three stages depending on intervention duration: a “pre-IRS” period during which the transmission in the local population reaches a stable state, followed by an “IRS” intervention period of ten or two years which reduces transmission rate (in what we call sustained vs. transient IRS, respectively), and a “post-IRS” period when transmission rates go back to their original levels only applicable to the transient IRS. Three different spatial configurations are implemented: a closed system with no migration, a semi-open system in which two individual parasite populations are connected via migration events, and a regionally-open system in which migrant genomes are sourced from a regional pool of *var* genes (Materials and Methods). The value of the baseline migration rate is inferred from field data from northern Ghana (Materials and Methods). Individual infections are only tracked locally, and other major stochastic events besides transmission include mutation of genes, recombination within and between genomes, and acquisition and loss of specific immune memory in hosts. (**B**) 14 different scenarios are considered in the simulation, indicated by Roman Numerals I to XIV. High migration rate is an order higher than the inferred baseline rate. Medium size regional pool and large one correspond to the aggregated diversity of a few and more than 10 individual local parasite populations with the region respectively. (**C**) Transmission intensity or effective contact rate (Materials and Methods) varies as a function of time, from pre-, to during, to post-intervention for a seasonal and closed system, with the two panels illustrating the sustained vs. transient implementation for the different reduction levels. (**D**) Different levels of intervention coverage are considered that reduce transmission rate log-linearly along the gradient of transmission intensity. For illustration purposes, we show here levels for a seasonal and closed system under sustained interventions. The lowest and highest reductions of the transmission rate correspond respectively to ∼20% and ∼90%, with colors corresponding to those in the left panel of C the sustained interventions. The highest reduction for transient interventions can correspond to more than 90%, reaching 96% for certain scenarios. (**E**) The study design of the three-round transient IRS in Bongo District in northern Ghana.

**Fig. 2.**
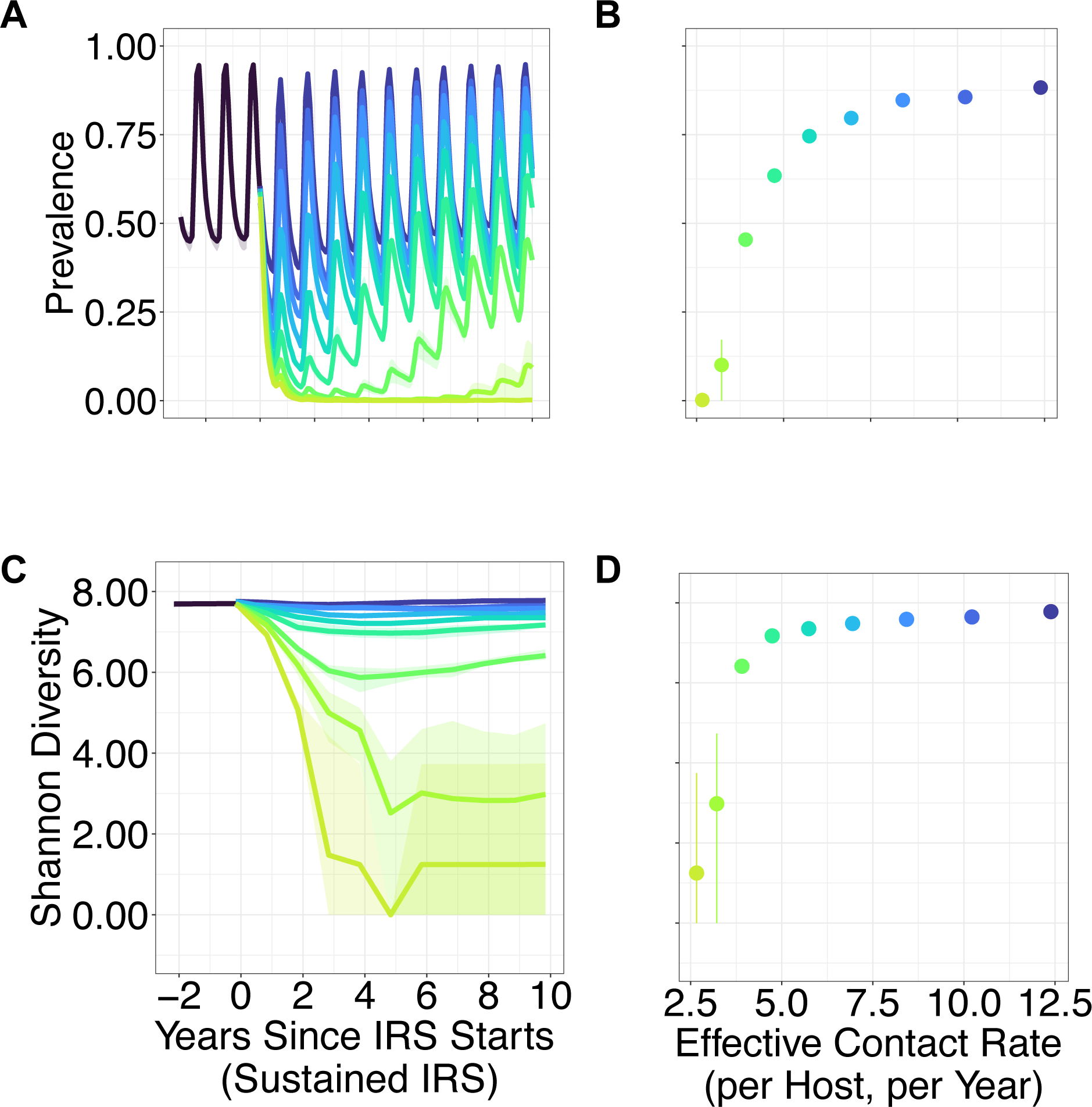
A seasonal and regionally-open system’s response to a series of sustained interventions (scenario VII in Fig. 1B) (**A**) Rebound dynamics of prevalence across time. The dynamics can be categorized into three regimes described respectively as fast rebound, transition, and slow rebound, depending on the behavior of prevalence over time. As the name indicates, for the fast-rebound regime, prevalence returns quickly to similar levels as those pre-intervention, whereas for the slow-rebound regime, prevalence does not recover within a decade and remain substantially lower, and the transition regime exhibits an intermediate recovery. (**B**) The long-term rebound prevalence levels at the end of the decade (sampled at the end of wet/high-transmission season) are shown against the transmission rates corresponding to the different intervention levels (with colors corresponding to those in Fig. 1D). (**C**) Rebound dynamics of *var* gene diversity measured by the Shannon Diversity Index across time. (**D**) Long-term rebound dynamics of *var* gene diversity against transmission rates (with values shown at the end of the decade). Both prevalence and *var* gene diversity exhibit a non-linear pattern against transmission rates. The fast-rebound regime corresponds to a wide interval, whereas the transition and slow-rebound regimes cover a narrow interval along the gradient of transmission rates. For the rebound dynamics of additional combinations of parameter values for closed, semi-open and regionally-open systems, see figs. S1 and S3.

The prevalence level reached after the rebound (a decade into sustained IRS, sampled at the end of the wet/high-transmission season for seasonal transmission) is highly nonlinear along the gradient of transmission intensity (Fig. 2B, figs. S1B). The fast-rebound regime occurs for a broad range within the gradient of transmission intensity, whereas the transition and slow-rebound regimes correspond instead to narrow ranges. Thus, the perturbed system experiences a sharp transition along the gradient of transmission intensities, from a rapid recovery down to a lingering low prevalence. Concomitantly, *var* diversity itself, whether measured as richness or the Shannon Diversity Index, also exhibits the three general behaviors, from the maintenance of high values, through the establishment of intermediate ones, to the eventual persistence of low levels, across the series of sustained IRS interventions (Fig. 2C, figs. S2A, figs. S3A). Post-intervention *var* diversity levels (a decade into sustained IRS) are also highly nonlinear with intervention transmission intensity (Fig. 2D, figs. S2B, figs. S3B).

The nonlinearity of the rebound in prevalence and corresponding *var* diversity levels along the gradient of transmission intensity holds, regardless of whether the system is under constant or seasonal transmission, and whether it is closed, semi-open, or regionally-open, over wide ranges of parameters (Fig. 2, figs. S1-3).

Although to examine long-term rebound dynamics we sustain the vector reduction for up to a decade in the numerical simulations, we find that the perturbed system in the slow-rebound regime can be prone to extinction at a faster time scale. Replicate runs can go extinct as early as in the fourth year into intervention (Fig. 3A), a period that is more comparable to what can be realistically implemented in empirical settings.

**Fig. 3.**
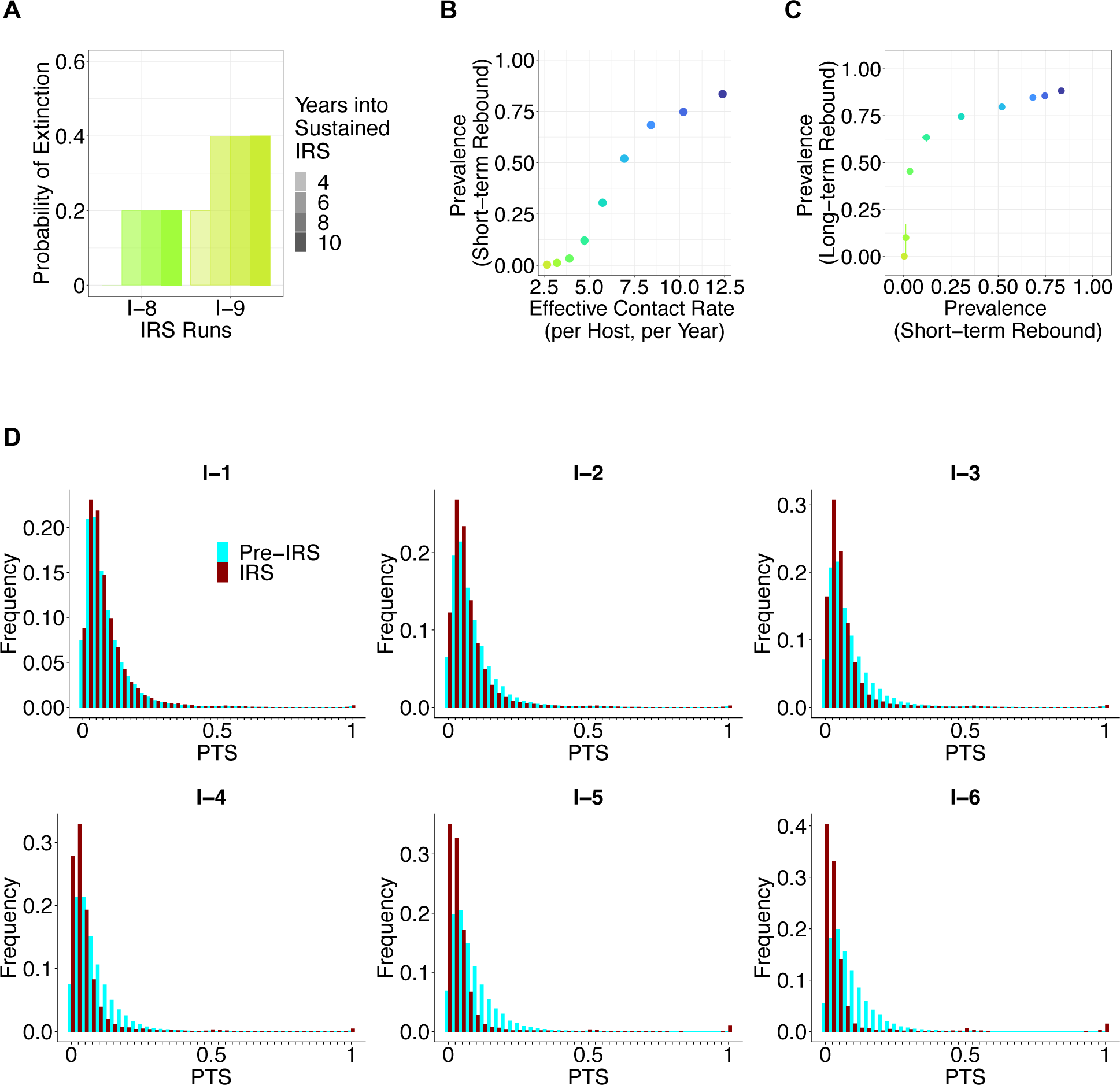
The probability of extinction in the slow-rebound regime, short-term vs. long-term rebound prevalence levels, and the population-wise pairwise type sharing or PTS distribution before and early into sustained interventions for a seasonal and regionally-open system (scenario VII in Fig. 1B) (**A**) Probability of parasite extinction calculated from replicate runs for intervention intensities representative of those in the slow-rebound regime. Extinction occurs as early as in the fourth year into sustained interventions. The two intervention levels correspond to those in Fig. 1D. (**B**) The short-term rebound prevalence levels at the end of the 2nd year of sustained intervention (sampled at the end of wet/high-transmission season) are shown against the transmission rates corresponding to the different intervention levels. For additional scenarios simulated, see figs. S5A. (**C**) Short-term rebound prevalence levels (in the second year of intervention, sampled at the end of wet/high-transmission season) are not good predictors of those in the longer term (at the end of the decade, sampled at the end of wet/high-transmission season). For additional scenarios simulated, see figs. S5B. (**D**) The pairwise type sharing or PTS distribution during sustained intervention without measurement error. With increased intervention intensities, and as the system approaches the transition regime, the PTS distribution exhibits the emergence of a second mode at complete overlap due to clonal expansion, and a shift toward lower values (of zero) at the lowest end due to reduction in meiotic recombination (or out-crossing) and hence the circulation of non-related strains. The emergence of the second mode is sensitive to the specification of parameters and disappears under measurement error (figs. S6-7). Thus, it is not a robust indicator, unsuitable for practical surveillance purposes, of approach to the transition regime.

Under low or mid-coverage vector control, transmission intensity is maintained at a somewhat high level (Fig. 1D). Immediately following intervention, the majority, or a high fraction, of infections can still get transmitted, resulting in a high surviving diversity (Fig. 2A, 2C, figs. S1A, S2A, S3A). Hosts’ immune profiles are heterogeneous because they were exposed to different subsets of the large circulating diversity before intervention. The high surviving diversity, coupled with hosts’ heterogeneous immune profiles, allows for a heterogeneous distribution of infection duration (figs. S4). Infections with longer duration are selected due to a reduction in transmission intensity. Thus, the distribution of infection duration immediately following intervention shifts to longer values compared to those before intervention (figs. S4). The selection for infections of longer duration effectively compensates for the reduction in transmission, allowing prevalence to start rebounding. Further into intervention, the loss of immunity due to the reduction in transmission and exposure increases individual susceptibility and consequently infection duration. Prevalence rebounds further, eventually reaching comparable levels to those pre-intervention. In contrast, under high-coverage vector control, immediately following intervention, transmission intensity and circulating diversity both drop to very low levels with a low fraction of infections getting transmitted (Fig. 1D, 2A, 2C, figs. S1A, S2A, S3A). Few infections can survive longer than the average waiting time between consecutive transmission events. The reduction in transmission intensity cannot be effectively compensated for by the shift in infection duration distribution towards longer values. Thus, the transmission system remains at low prevalence and diversity.

Having documented a threshold behavior of the transmission system in its response to intervention, we address next the existence of indicators revealing its proximity to the transition regime. Because prevalence is a commonly tracked quantity in malaria surveillance, we first examine whether it can function as one such indicator. The short-term rebound prevalence levels following simulated sustained IRS (two years into intervention) show a much more linear behavior with intervention levels than those after longer sustained control (Fig. 3B, figs. S5A). The relationship between these two types of prevalence levels is non-linear and the degree of nonlinearity varies across different spatial configurations and parameter values (Materials and Methods, Fig. 3C, figs. S5B). Overall, short-term prevalence levels are only weakly correlated with, and are by themselves only poorly predictive of, long-term rebound prevalence levels. Moreover, high-quality prevalence data is often unavailable in empirical settings, and efforts to collect these data can be hindered by low testing rates and imperfect detective power for asymptomatic infections (*40*). Thus, prevalence by itself does not provide sufficient information on either the proximity of the perturbed system to the transition regime or the loss of resilience of the epidemiological system.

As complementary information, we therefore address indicators related to the structure of diversity from the perspective of *var* genes and isolates. As we seek to apply such indicators to field data, we also consider whether they are robust to measurement errors common in sampling schemes (Materials and Methods). We further focus on responsive indicators that rely on sampling immediately following the start of intervention, specifically two years into it (at the end of wet/high-transmission season for seasonal systems).

We first compared whether the distributions of pairwise type sharing (PTS) differ between before and early into intervention (*38*). PTS, a similarity index analogous to the Sørensen or Jaccard Index (*41, 42*), describes the fraction of shared DBLα types between any two isolates and hence quantifies the repertoire similarity of parasites circulating in host population. If each isolate consisted of a monoclonal infection by a repertoire of one *var* gene, sample PTS would be equivalent to homozygosity, and the comparison of PTS within and between populations would be equivalent to Wright’s fixation indices. PTS can therefore be viewed as a simple extension of fixation indices to the case where multigene family members of unknown allelic status have been collected (*38, 43*). A second mode at complete overlap emerges in the PTS distribution when the perturbed system approaches the transition regime (Fig. 3D). Concomitant with the appearance of the high overlap mode, the low end of the PTS distribution shifts towards zero or extremely low values (Fig. 3D, figs. S6-7) due to reduced outcrossing. Outcrossing creates relatedness and increases similarity between parental and offspring strains. Because the frequency of outcrossing events scales with transmission intensity, intervention results in a decrease in outcrossing rates and an increase in clonal transmission of non-related strains. Therefore, the PTS distribution is pulled into the two opposite extremes of complete and no, or extremely low, overlap. The emergence of this second mode at complete overlap is however sensitive to parameter specification and measurement error. It specifically becomes less apparent when the circulating diversity is the highest among all the simulated scenarios (Materials and Methods), effectively disappearing when we do not completely sample all the *var* genes constituting infections (figs. S6-7). This happens because this incipient appearance of clonality involves too few parasites in the population, a result of intervening in a high-transmission endemic system where the original diversity is high. Any subset of strains from this original diversity which survived intervention, will likely be dissimilar from each other. Unless transmission remains sufficiently low with importation of *var* gene diversity curtailed so that clonal transmission follows a bottleneck in strain diversity, the second mode in the PTS distribution will be insignificant. In addition, this second mode of complete overlap becomes more diffused under measurement error: two identical strains will have a high but uncertain PTS value within some range, depending on which subset of their *var* genes are sequenced and typed. Thus, the appearance of the second mode cannot be reliably used for empirical surveillance.

Instead of the population-wise PTS, we then examine the age group differences at the low-end of the PTS distributions. A previous study has shown age-dependent patterns of PTS in empirical data, with infections in children (1-10 years old) exhibiting more related *var* repertoires compared to adults (>20 years old), consistent with the effect of immune selection (negative frequency-dependent selection) (*24*). We therefore compare these distributions in children and adults from the simulation output across the series of sustained transmission-reducing interventions. We find a reduced difference as the system moves from the fast-rebound regime toward the transition one. Specifically, the PTS distribution within children manifests a mode around zero or extremely low overlap, which was originally present in the PTS distribution of adults. In other words, the PTS distribution between the two age groups becomes more similar, overlapping at the lowest mode when the system approaches and enters the transition regime (Fig. 4A, figs. S8-21A). This behavior is robust to sampling schemes simulating the collection of field data (Materials and Methods).

**Fig. 4.**
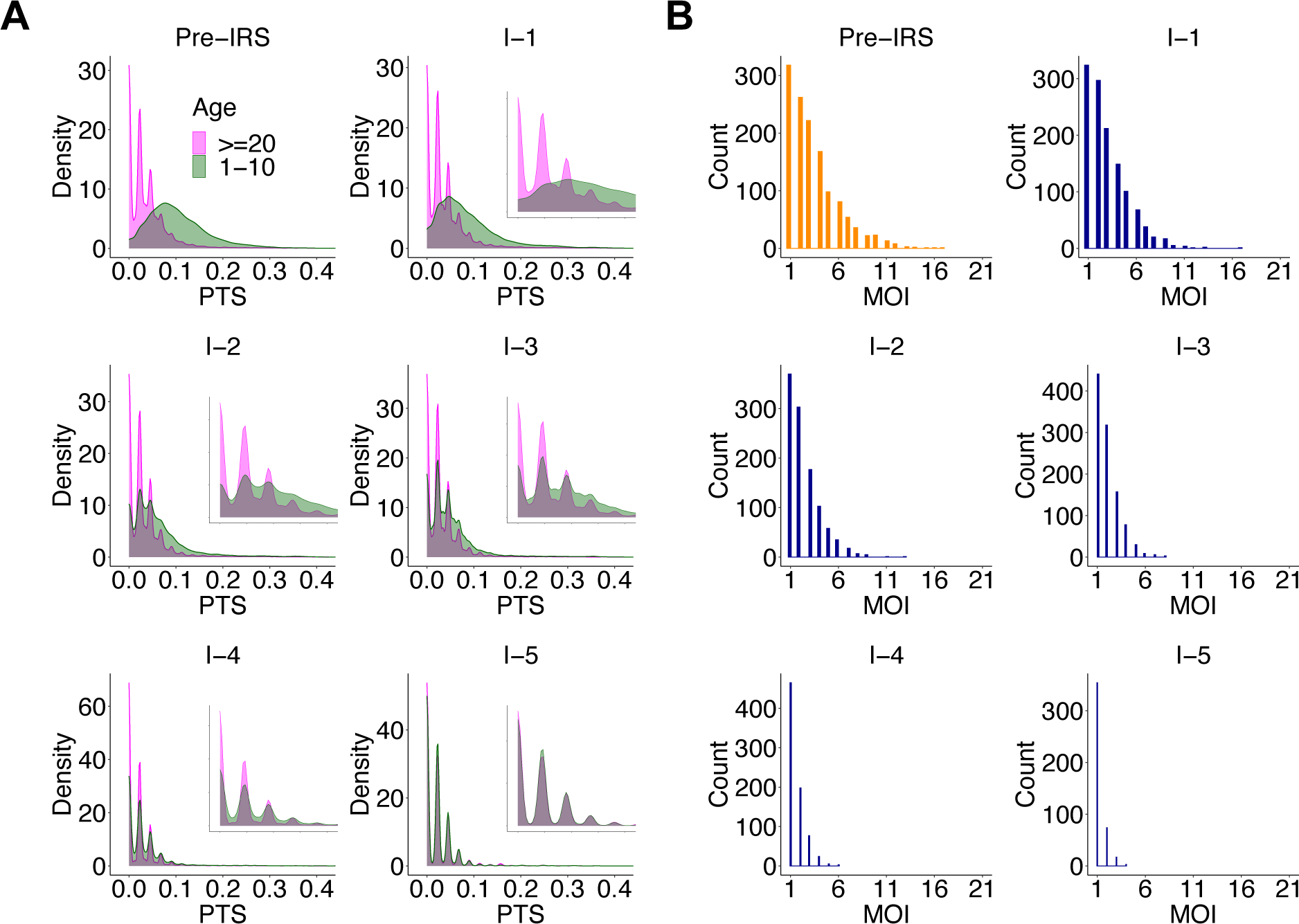
Molecular indicators of the proximity of the perturbed local system to the transition regime for a seasonal regionally-open system with a medium-size pool and a baseline migration rate (scenario VII in Fig. 1B) (A) As the system approaches the transition regime, the difference between PTS distributions across age groups is significantly reduced and even disappears. In particular, the two distributions almost completely overlap at the lowest mode around 0. The x-axis range for inset figures is 0-0.1. (**B**) MOI distribution starts to center around 1 and 2 with the majority of infections being either mono-clonal or multi-genomic with two genetically distinct parasites. These two molecular indicators are robust under implemented sampling schemes and sampling limitations representative of those encountered in the collection of field data (figs. S8). Molecular indicators for regionally-open systems with other setups or closed and semi-open systems can be found in figs. S9-21.

A second candidate for an indicator quantity is the distribution of the multiplicity of infection (MOI), defined as the number of genetically distinct parasite strains co-infecting a single human host (*33, 44, 45*). MOI, also referred to as complexity of infection (COI), is one of the most frequently reported proxies for parasite transmission (*46, 47*). Although a variety of statistical approaches have been developed that rely on various sources of genotypic data, the estimation of MOI remains challenging for high-transmission endemic settings where individuals typically carry multiple co-occurring infections (i.e., MOI > 1) (*48*). The low to non-existent overlap of *var* repertoires enables estimation of MOI on the basis of the number of *var* genes sequenced from an individual’s isolate (*24, 35*). This so-called “*var*coding” method has been shown to provide accurate estimation in numerically simulated infections, and to perform better when transmission is high than a previous method based on a collection of neutral single nucleotide polymorphism loci (SNPs) (*48*). Having extended this approach to a Bayesian framework to account for measurement error in the sequencing (*21*), we apply the approach here to derive MOI estimates at the population level across the series of IRS interventions. We find that the MOI distribution becomes increasingly centered at low values around one and two, as the system approaches the transition regime (Fig. 4B, figs. S8-21B). Specifically, whereas for the fast-rebound regime the majority of infections remain multi-genomic (MOI > 1), the fraction of multi-genomic infections decreases significantly when the intervened system approaches the transition regime. For the transition and slow-rebound regime, the majority of infections become mono-genomic (MOI = 1). This indicator is also robust under sampling schemes simulating the collection of field data (Materials and Methods).

We apply next the above indicators to an interrupted time-series study involving three age-stratified cross-sectional surveys undertaken at the end of the wet/high-transmission season from Bongo District in northern Ghana, respectively before, during, and immediately after the transient IRS intervention (Fig. 1E) (*21, 35*). Following IRS, the difference in the PTS distribution across the two age groups is reduced, almost completely overlapping at the lowest mode (Fig. 5A), and the MOI distribution shifts towards a high proportion of mono-genomic infections or multi-genomic ones with only two genetically distinct parasites (Fig. 5B). These patterns suggest that intervention had pushed the empirical system near the transition regime. Similar patterns are found for the survey immediately after the IRS intervention (termed as the post-IRS phase, Fig. 1E), suggesting the impact of IRS persisted through the wet/high-transmission season after it was discontinued.

**Fig. 5.**
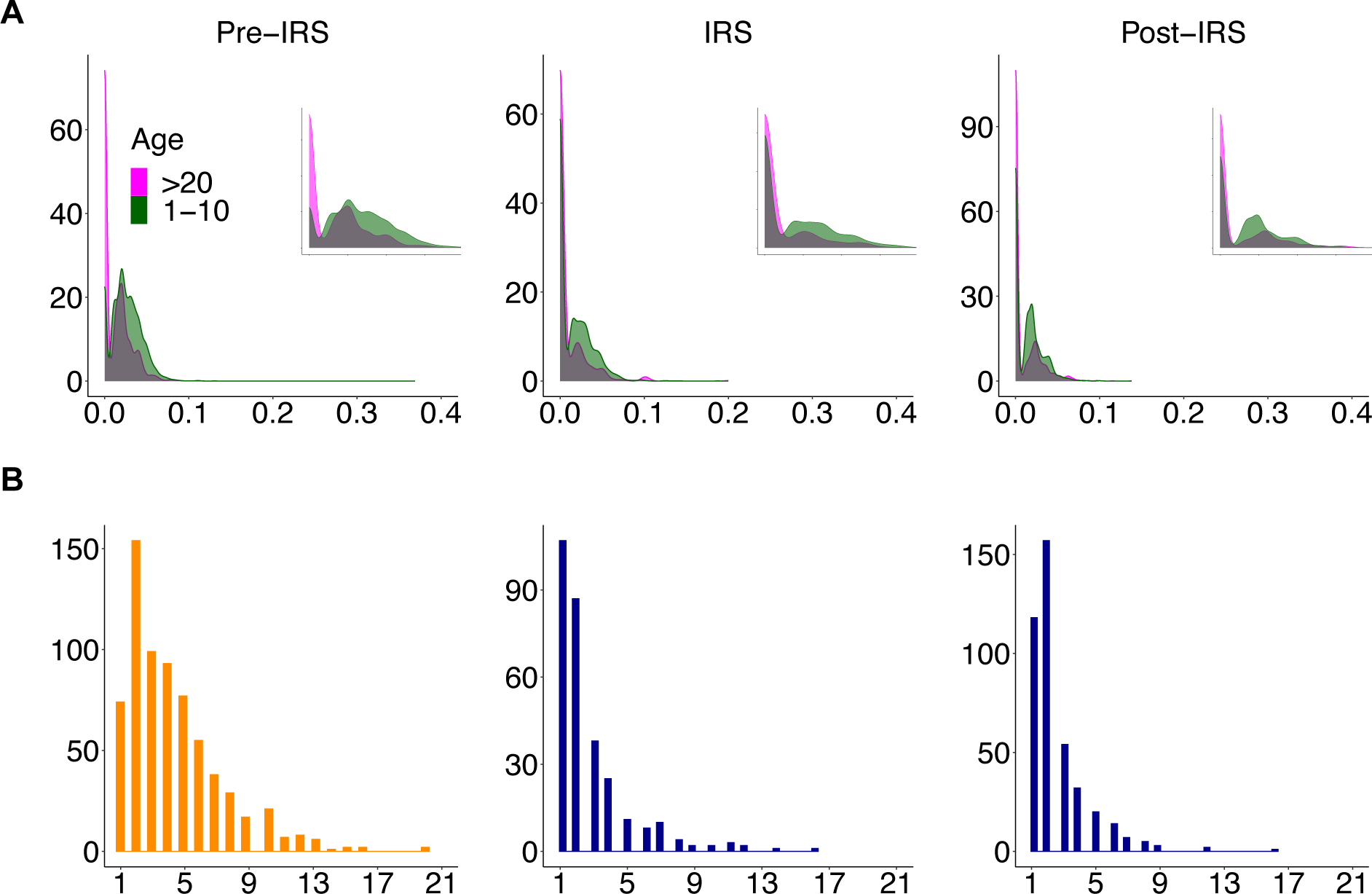
The two molecular indicators applied to the field data from northern Ghana before, during, and immediately after the three-round transient IRS intervention, indicate proximity to the transition region where rebound prevalence decreases sharply with intervention intensity. (**A**) The PTS distribution between the two age groups becomes more similar, especially for its lowest mode, which is almost completely overlapping. The x-axis range for inset figures is 0-0.075. (**B**) The MOI distribution exhibits its main mode at one, with the majority of infections becoming either mono-clonal or multi-genomic with two genetically distinct parasites. Those patterns persisted during the year right after when the IRS was discontinued (the post-IRS panel), although to a slightly lesser degree.

Because the three-round IRS intervention was only implemented over a two-year window, we return to the computational model to examine a series of transient interventions of this length across the different spatial configurations (Fig. 1C-D, Materials and Methods). Overall, the simulated systems are highly resilient to transient interventions, especially when embedded within a regional pool. After interventions are discontinued, regionally-open systems always rebound rapidly in prevalence back to pre-IRS levels, regardless of IRS coverage and configuration of their regional pool (Fig. 6A, figs. S22). The majority of closed and semi-open systems rebound rapidly as well. However, in a few instances, prevalence remains confined to intermediate or low prevalence levels requiring more than a decade to fully rebound (Fig. 6A, figs. S22). In these, IRS coverage had been at the highest end and sufficiently strong to bring prevalence levels and parasite population sizes close to zero (Fig. 6A, figs. S22), with a low *var* diversity during IRS (Fig. 6C).

**Fig. 6.**
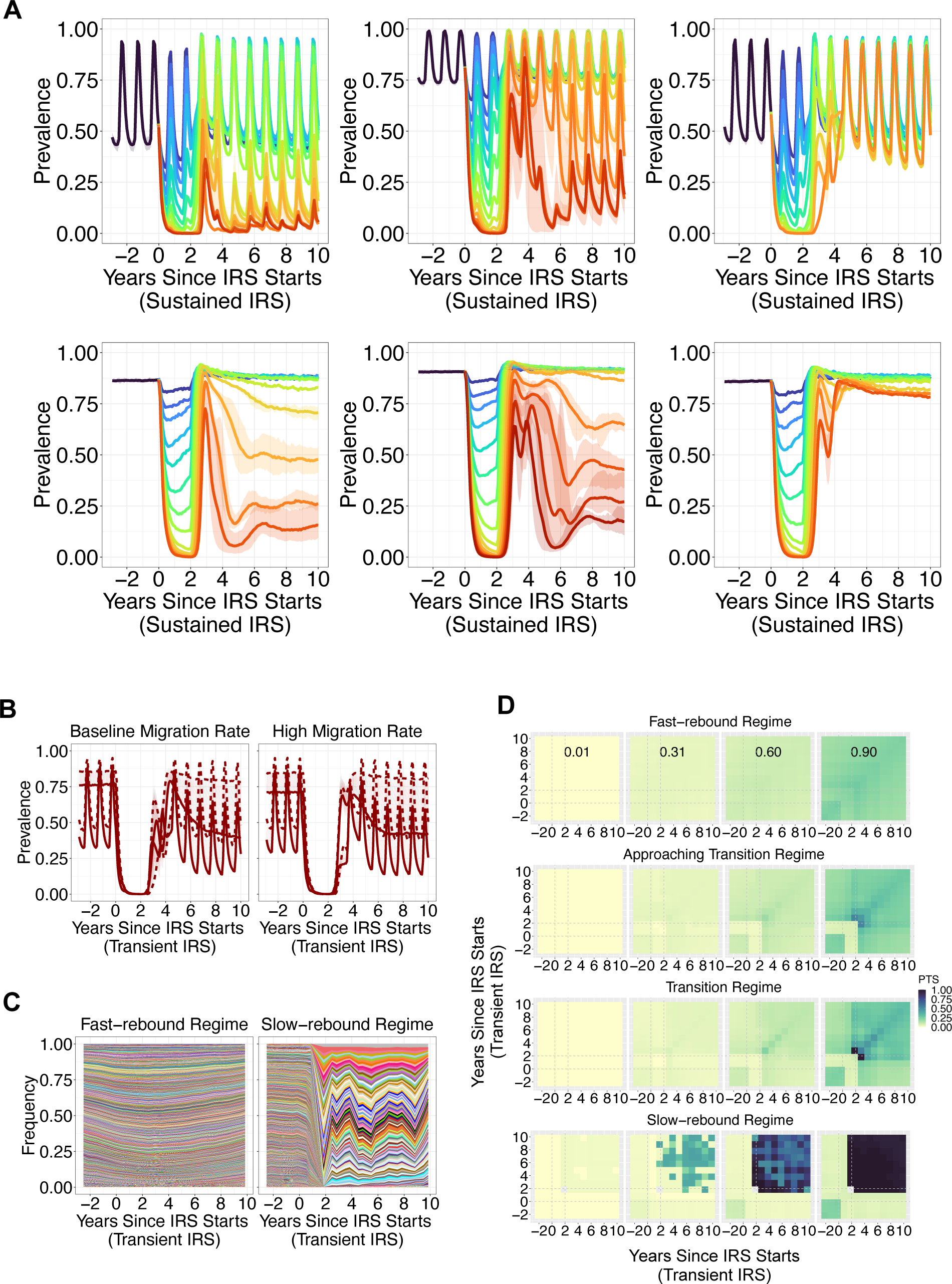
The rebound dynamics in prevalence across time, under a series of transient interventions (2 years); the diversity of migrant genomes and the degree of novelty they introduce to the local parasite population vs. rebound dynamics to transient intervention for a seasonal regionally-open system; the parasite genetic structure of closed systems in the fast rebound, transition, and slow rebound regimes to transient intervention. (**A**): For closed and semi-open systems, the dynamics can be categorized into three regimes described respectively as fast rebound, transition, and slow rebound, depending on the behavior of prevalence over time. For regionally-open systems, the dynamics are always those of fast rebound kind. Such resilience is enabled primarily by the effective importation of novelty post interventions. The top panel from left to right: seasonal and closed systems (scenario I in Fig. 1B), seasonal and semi-open systems with a baseline migration rate (scenario III in Fig. 1B), seasonal and regionally-open systems with a medium-size pool and a baseline migration rate (scenario VII in Fig. 1B). The bottom panel from left to right: non-seasonal systems, corresponding to scenario II, V, XI in Fig. 1B. Simulation runs with other parameter values are shown in figs. S22. (**B**): The rebound dynamics of regionally-open systems with a medium-size pool to transient (2 years) and highest-coverage interventions. These systems receive migration from either a dynamic regional pool with regular substitution of common genes in that pool, hence effective importation of novelty via migration events, or a static regional pool which does not update its gene composition. Fluctuating lines correspond to seasonal transmission whereas the relatively non-fluctuating lines represent constant transmission. Dashed lines correspond to dynamic regional pools whereas solid lines correspond to static ones. The left panel assumes a baseline migration rate whereas the right panel assumes a migration rate that is an order higher than the baseline one (Materials and Methods). Rapid rebound occurs when the pool is dynamic, and the importation of novelty is effective. (**C**): The frequency of genes circulating in the host population varies as a function of time, from pre-, to during, to post-intervention. Each color in the stacked area plots refers to a different gene. Shown here are two instances of seasonal and closed systems under transient interventions (scenario I in Fig. 1B). The left panel corresponds to a perturbed system which demonstrates a fast rebound dynamics (circulating genes are plentiful and typically exhibit low frequency), whereas the right panel corresponds to a system which demonstrates a slow-rebound dynamics (circulating genes are low in numbers with each at high frequency). (**D**): When the system rebounds from a population bottleneck induced by transient interventions (2 years), in the fast-rebound regime, clonal expansions are limited and circulating strains are maintained to be dissimilar from each other, measured by PTS distributions. The representative four quantiles of these distributions are at low values, indicating that strains sampled at the same times and those between present and past (diagonal versus off-diagonal values, respectively), are dissimilar. In contrast, circulating strains are highly similar in the slow-rebound regime. Clonal expansion dominates.

We further investigate why the regionally-open systems are highly resilient to transient intervention. Because antigenic diversity facilitates immune evasion and further infection, we address the question from the perspective of change in *var* diversity and associated strain structure by examining how the diversity of migrant genomes and the degree of novelty they introduce to the local parasite population impact the resilience of a regionally-open system. We specifically implement the following two scenarios: a high migration rate per se, and one that is additionally accompanied by a highly diverse gene composition of migrant genomes. We consider for this purpose a static regional pool of medium size (lower than the highest value considered for high-transmission endemic regions) that does not update its genes, versus a dynamic one of the same size that does so continuously. Migrant genomes drawn from this static regional pool necessarily import the same set of *var* genes repeatedly to the local parasite population, and therefore hosts typically develop more complete immunity against their products and the system rebounds in prevalence, although not rapidly. In contrast, migrant genomes drawn from the dynamic regional pool import new *var* genes to which hosts are susceptible, and the system rebounds in prevalence rapidly (Fig. 6B). The transient intervention considered here is among the highest-coverage ones, reducing prevalence and parasite population size close to zero, and *var* diversity to very low values. We do not repeat this analysis for regional pools of the largest size because they necessarily entail migrant genomes introducing a high degree of novelty to the local parasite population, regardless of the rate of update of their gene composition. Regionally-open systems are highly resilient to transient intervention because migrant genomes effectively introduce novelty and help the recovery in *var* diversity once control is discontinued.

In contrast, closed and semi-open systems can lose their resilience under transient high-coverage intervention. We examine how the disruption of parasite genetic structure typical of high transmission underlies this loss of resilience. For closed and semi-open systems that exhibit a fast rebound, their parasite populations remain large and retain *var* gene diversity characteristic of high-transmission regions during transient intervention (Fig. 6C-D). After control is discontinued, surviving parasite populations expand, and the degree of clonal transmission is low. Circulating genes are plentiful and typically exhibit low frequency (Fig. 6C), and strains are dissimilar from one another as reflected in PTS distributions. These distributions capture the degree of overlap of isolates that are circulating within the same time period as well as those in the past and present (diagonal and off-diagonal values, respectively, in Fig. 6D). Their four quantiles systematically remain low. Hosts’ exposure to dissimilar strains facilitates immune evasion and further infection, which in turn promotes the generation and maintenance of *var* gene diversity.

For closed and semi-open systems that exhibit a transition or slow rebound, their parasite populations approach zero with low *var* diversity during transient intervention (Fig. 6C). After control is discontinued, small surviving parasite populations expand, resulting in a high degree of clonal transmission. Circulating genes are low in numbers with each at high frequency (Fig. 6C) and strains are similar to one another (Fig. 6D). Hosts’ exposure to highly similar strains throughout time compromises immune evasion and further infection, which in turn reduces the generation and maintenance of *var* gene diversity. In our simulations, such systems require more than a decade to recover their prevalence fully.

## DISCUSSION

We have described a non-linear and sharp response to the intensity of a sustained transmission-reducing intervention, with an agent-based model that explicitly tracks specific immune memory to strain variation encoded by a hyper-variable multigene family. The perturbed transmission system experiences a threshold behavior, losing its resilience when pushed into a narrow transition region toward a regime where prevalence rebounds only slowly with an associated extinction risk of the parasite. By examining the relationship between the loss of *var* diversity and the disruption in the strain structure of limiting overlap, we have identified molecular indicators providing information on the approach of the perturbed transmission system to the transition between a fast and slow rebound to high prevalence. These indicators rely on sampling following intervention, effectively complementing the monitoring of the transmission system through epidemiological quantities over time. They can inform timely adjustment of intervention efforts, to push the system into the transition regime and beyond, into the slow-rebound behavior enhancing local parasite extinction, to do so more rapidly than by observing the temporal trajectory of prevalence itself.

The response behavior we have described involves a clear coupling between the rebound dynamics in prevalence and those of *var* gene diversity. In the fast-rebound regime, the heterogeneity of hosts’ immunity profiles and large surviving strain diversity underlie a heterogeneous distribution of infection duration. Immunity loss increases hosts’ susceptibility to infection and contributes to further lengthening infection duration. Selection for infections with longer duration in response to reduced transmission, effectively compensates for this reduction. In contrast, in the slow-rebound regime, few infections can survive for sufficiently long times to withstand the waiting time between consecutive bite events, resulting in a low surviving diversity and population size of the parasite. Selection for infections with longer duration can no longer effectively compensate for the reduction in transmission.

We illustrated the application of the identified indicators to field data from Bongo District in northern Ghana (*21*). Their patterns suggest that the transmission system had been brought close to transition by the two-year transient intervention, and that therefore sustaining and intensifying these efforts would have been worthwhile.

Similar patterns for the two molecular indicators of approaching the transition phase persisted through the wet/high-transmission season after the three-round transient IRS was discontinued. However, prevalence and the population genetics of the *var* multigene family rebounded rapidly, already returning to their pre-IRS characteristics when surveyed 32-month after IRS was discontinued (*21*). During the transient intervention, despite a considerable reduction in parasite prevalence (by ∼40-50% compared to the pre-IRS level), estimates of parasite population size remained large and *var* population genetics showed persistent characteristics of high transmission regions (*21*). These characteristics would provide a large enough founding population to effectively maintain a low degree of clonal transmission after IRS was discontinued. The local parasite population was likely embedded in a large regional pool, which enables the maintenance and rapid recovery of *var* diversity. Resulting immune evasion and associated transmission would have further promoted the generation and maintenance of diversity, enabling a positive feedback between transmission and diversity which has been previously described in two other malaria models (*49, 50*). The relationship between the response of the transmission system described here in a stochastic individual-based model and the tipping point behavior identified in a deterministic compartmental population model of malaria (*50*) should be investigated.

Although we relied initially on relatively long interventions (i.e., a decade), we also examined more transient ones, which underscored the fundamental importance of spatial context through the importation of *var* gene diversity. Although regionally-open systems not surprisingly always exhibit a fast-rebound dynamics in prevalence regardless of intervention coverage, semi-open and closed systems can experience the full range of possible temporal trajectories. The positive feedback described above should remain highly efficient for regionally-open systems, or closed and semi-open systems with fast resurgence once transient intervention is discontinued. In this case, surviving parasite populations can rapidly expand, yet the degree of clonal transmission remains low. Circulating *var* diversity remains high and strains are dissimilar from one another despite a population bottleneck. In contrast, the positive feedback is weak for closed and semi-open systems which enter the transition and slow-rebound regimes. After control is discontinued, clonal expansion dominates, resulting in the circulation of highly similar strains. Thus, as for intervention in other transmission settings, coordinated regional efforts and information on sources and sinks are clearly important. Ways to infer spatial context and the importation of *var* gene diversity from genomic data should be investigated.

Our computational model did not consider generalized (VSA-transcending) immunity or functional divergence between genes (*4, 51, 52, 53, 54, 55, 56, 57, 58, 59, 59, 60, 61*). Future work will examine the performance of the identified indicators under further aspects of immunity and incorporate different *var* gene groups. We also used genetic variation as a proxy for antigenic variation (*38*) because a genotype to phenotype map is not available for *var* genes. Future work should examine ways to derive aspects of the map from a variety of perspectives, including bioinformatics, the fitting of ABMs, and protein structure prediction based on deep learning. For regionally-open systems, our simulation considers a local parasite population embedded within a regional pool providing the source of migrant genomes and does not represent metapopulation dynamics explicitly. An explicit simulation of multiple parasite populations in our ABM is computationally expensive. An open population with continuous migration from a regional pool is an alternative typical setting of many assembly models in community ecology.

The World Health Organization has recently underscored the importance of surveillance for malaria intervention (*62, 63*). Molecular epidemiology provides approaches to complement traditional surveillance, with methods best suited for moderate-to low-transmission intensities (*64, 65, 66*). Deep sampling of sequences of *var* genes in host population provides a basis to complement these efforts for high transmission, with a focus on the high diversity that characterizes the *var* multigene family in these regions. Our results establish a link between the population dynamics of the disease and observations of parasite population genomics from the perspective of hyper-variable antigen-encoding genes. They indicate that the structure of diversity revealed via molecular surveillance can inform intervention against malaria in high-transmission endemic settings. These findings should enhance our understanding of transmission dynamics where large standing pathogen diversity represents a major challenge to control efforts.

## MATERIALS AND METHODS

### Stochastic agent-based model

We extend a previous computational model (*29*) whose key assumptions and processes are described in supplementary Materials and Methods. Model parameters and symbols are listed and explained in Data file S1. The model is an agent-based (individual-based), discrete-event, and continuous-time stochastic system in which all known possible future events are stored in a single event queue along with their putative times, which may be fixed or drawn from a probability distribution with a certain rate. We choose values for the rates of these events based on the literature of malaria epidemiology, related field studies, and *in vitro* or *in vivo* values (see supplementary Materials and Methods and Data file S1 for further details). When an event happens, it can cause the addition or removal of future events in the queue, or the modification of their rates, resulting in a recalculation of putative times. This approach is implemented with the next-reaction method (*67*), which optimizes the Gillespie first-reaction method (*68*).

Individual infections and the immune history of individual human hosts are tracked, and evolutionary mechanisms including mitotic/ectopic recombination and mutation are modeled explicitly. We simulate a local host population of a given size which can assume different spatial configurations (see section “Seasonality and spatial configuration of the local transmission system” below). At the beginning of each simulation, a small number of hosts are randomly selected and infected with distinct parasite genomes from the random assembly of *var* genes from a “regional” pool to initialize local transmission.

Each parasite genome is represented as a specific combination of 45 (non-upsA) *var* genes, and each *var* gene is considered a linear combination of two epitopes (alleles) based on the empirical description of two hypervariable regions in the *var* tag region of the DBLα domain (*54*). Mosquito vectors are not explicitly represented as agents in the model. Instead, we consider an effective contact rate (hereafter, the transmission rate, which under some assumptions is effectively equivalent to vectorial capacity) which determines the times of local transmission events. Donor and recipient hosts are selected at random, and transmission occurs if the former carries active blood-stage infections, and the latter has not reached the carrying capacity of its liver-stage. We model ectopic recombination among genes within the same genome during the asexual stage, and meiotic recombination between strains during sexual replication inside the mosquito vector. The expression of genes in the repertoire is sequential and the infection ends when the whole repertoire is depleted. Hosts acquire transitory immune memory toward the product of expressed epitopes, and such memory precludes expression of such epitopes in future infections. The total duration of infection of a particular repertoire is therefore proportional to the number of unseen epitopes by the infected host across all individual *var* genes of a given repertoire. See supplementary Materials and Methods for further details of the agent-based model (ABM).

Extensions of the original computational model (*29*) implemented for this work concern the regionally-open spatial configuration, ectopic recombination events, and within-host dynamics. In particular, the regional pool for the regionally-open system now updates its genes at a rate reflecting the substitution rate of genes in individual local parasite populations within the region. Ectopic recombination events can now generate truly novel alleles instead of simply shuffling the alleles of the two parental genes. The current formulation of the model further allows ectopic recombination events to be associated with a load such that there is a certain probability of generating non-functional offspring genes which replace functional parental genes. These non-functional genes will not be expressed and hence reduce infection duration of repertoires within infected hosts.

### Seasonality and spatial configuration of the local transmission system

We simulate both constant and seasonal transmission dynamics. We implemented seasonality by multiplying a scaling constant by a temporal vector of 360 days, containing the daily number of mosquitoes over a full year. In other words, the temporal vector represents the mosquito abundance, and the scaling constant encapsulates all other parameters involved in vectorial capacity. The product of both gives the effective contact rate. To obtain this temporal vector, (*79*) used a deterministic model for mosquito population dynamics from (*80*). The model was originally developed for *Anopheles gambiae* and consists of a set of ODEs describing the dynamics of 4 mosquito stages: eggs, larvae, pupae, and adults (*80*). Seasonality is implemented via density dependence at the egg and larva stages as a function of rainfall (availability of breeding sites). We adopt either constant effective contact rates or seasonal ones, which result in EIR or FOI values typical of high-transmission endemic regions (*75*) (see section “Repertoire transmission” and “Within-host dynamics” in supplementary Materials and Methods).

We simulate transmission dynamics for closed, semi-open, and regionally-open systems. For closed systems, after the initial seeding of local transmission from a pool of *var* genes, migration is discontinued. Thus, *var* diversity is generated intrinsically by the dynamics of the ABM for the local population. For semi-open systems, two individual populations coupled via migration are explicitly simulated. They represent empirical transmission systems which are mainly coupled to one or a few neighboring sites. Regionally-open systems represent a local transmission system that is connected with a few to many individual neighboring parasite populations within a broader regional scale. Instead of simulating those individual parasite populations explicitly, we let the focal population receive migration from a regional pool of *var* genes of a certain size. This regional pool acts as a proxy for regional parasite diversity, i.e., diversity of the aggregate of individual local parasite populations in the region. Because each parasite genome is a repertoire of a given number of *var* genes, migrant genomes are assembled from random sampling of *var* genes from the regional pool. Estimation of key parameters concerning the regional pool is described in the following sections.

### The temporal course of a simulation and the series of IRS interventions

Each simulation follows either two or three stages (Fig. 1A, 1C): (i) a pre-IRS period during which the local parasite community is assembled and the transmission system reaches a (semi-) stationary state before the intervention; (ii) an IRS period of either 10 or 2 years during which transmission is decreased (referred to as sustained and transient IRS, respectively); (iii) a post-IRS period when transmission rates recover to pre-IRS levels (only applicable to the transient IRS simulation, since the sustained one extends for the full decade).

We simulate a series of IRS intervention efforts, which reduce transmission rates to levels that are log-linear along the gradient of transmission intensity (Fig. 1D). The lowest and highest coverage of the IRS intervention efforts correspond to a reduction of 20% and more than 90%, respectively.

The IRS intervention efforts are assumed to be regional for open systems. In empirical settings, regional intervention efforts imply that similar efforts are applied to both the local parasite population of interest and the neighboring parasite populations which exchange genomes with the local population via either short-distance or long-range dispersal of mosquitoes or human hosts. All these parasite populations then experience a similar level of reduction in transmission and consequently in their prevalence. As a result, during IRS, the importation of migrant genomes from these neighboring locations also decreases. To implement regional interventions for regionally-open systems, we let migration rates be proportional to the local transmission rate and local prevalence level, both being surrogates of transmission rates and prevalence levels of neighboring populations.

We assume the size of the regional pool remains unaffected during and post intervention because the aggregate of *var* genes across individual parasite populations should remain high, even when intervention is regional and of high-coverage. We do so because this work uses the simulated “regionally-open systems” to represent high-transmission endemic regions in empirical settings, where immigration is known to represent an important challenge to control and elimination efforts. Here, regional interventions rarely eliminate the majority of individual parasite populations.

### Estimation of the baseline migration rate for the implementation of open systems

We rely on empirical data on *var* genes from Bongo District, a high-transmission region in northern Ghana to infer the rate of gene exchange between populations, and to obtain a reasonable estimate of migration rates to implement the open transmission systems. The fast-evolving *var* genes under immune selection provide a higher resolution than neutral loci for recent migration events. Following (*81*), we calculate Jost’s D (*82*), a measure of population divergence for highly diverse genetic markers, to quantify *var* gene differentiation within and between two proximal catchment areas (i.e., Vea/Gowrie and Soe) in Bongo District for which molecular sequences were previously obtained at the end of the wet/high-transmission season prior to the IRS intervention (*21, 24*). Furthermore, under the finite-island model with infinite alleles, the divergence between two populations, *D*, is proportional to the local innovation (primarily ectopic recombination) rate, and inversely proportional to the migration rate between the two populations. These relationships are intuitive because local innovation events generate new genes unique to the specific local population and hence promote genetic differentiation and isolation between populations, whereas migration events promote genetic exchange and hence reduce divergence between populations. We estimate *m*, the percentage of contacts leading to infection due to migration relative to local transmission events, by dividing the local innovation rate, i.e., the ectopic recombination rate of the *var* genes, by *D*. This approach was initially proposed for single-locus genes with multiple alleles (*82*). Here we apply it to the *var* multigene family, effectively treating it as if it were single-locus, and different variants of *var* as if they were different alleles of this locus. We consider a range of ectopic recombination rates representative of those in nature, although values of the ectopic recombination rate have been measured only *in vitro* (*26*). We consider the daily rate of ectopic recombination, and the percentage of migration rate per day relative to daily local transmission events. The estimated baseline migration rate is around 0.0026, which translates to a percentage of migrant contacts relative to local ones of around 0.26%. We simulate transmission dynamics for both semi-open and regionally-open systems with migration rates equal to, or an order of magnitude higher than, this estimated baseline.

### Estimation of the baseline rate at which genes are substituted in the regional pool

We estimate a value for the rate at which genes in the regional pool are substituted. We refer to this rate as the substitution rate hereafter. As the regional pool represents an aggregate of diversity circulating in individual local populations within the region, this rate can be approximated by the substitution rate of genes in individual local populations. Specifically, we can view a regional pool as the aggregate of diversity of a certain number of equally sized individual parasite populations. Note that we consider regional pools consisting of diversity in the form of the effective number of equifrequent genes and we do not specify a frequency distribution of genes in these regional pools. The collection of this effective number of equifrequent genes, in the ideal scenario, would exploit hosts’ immune space equivalently relative to the collection of genes at their original “true” frequency. We use the inverse Simpson diversity index (or, one over the expected homozygosity) (*82*), 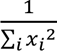, to determine the effective number of genes in individual local parasite populations of our simulated closed and semi-open systems, which is at the order of one to two thousand. Moreover, connected individual local parasite populations of these simulated semi-open systems share a significant fraction of their genes when ranked by frequency, around 50% or more. Therefore, we let the number of genes in the regional pool span from several thousand to over tens of thousands, to mimic from a few to about ten to twenty individual local parasite populations. We specifically consider two setups for the regional pool representing roughly the two extremes, referred to as “medium” versus “large” pool scenarios respectively.

The substitution rate of common genes in a local population depends on the local innovation rate, i.e., the rate at which new variants are generated from ectopic recombination and mutation, and the fixation probability, or the invasion probability, of these new variants (*84, 85*). Fixation of a new gene variant in this context refers to a single copy of the variant present at a given time point having descendants in the population after a very large amount of time has passed, which typically entails that the frequency of the new variant reaches a certain threshold and becomes sufficiently common so that it is much less susceptible to genetic drift. When the system is at, or close to, a stationary state, the fixation of a new gene variant would imply the loss or replacement of an older common gene variant, hence we term this process and its rate as substitution of genes and substitution rate respectively. The rate of ectopic recombination and mutation has been measured *in vitro* (*26*). The magnitude of the fixation or invasion probability can be derived on the basis of population genetic models and the simulation output from our ABM. From the two together, we can obtain an expectation for the substitution rate as follows.

We write the invasion probability of new variants, or genes, or alleles in the system. At the stationary state, malaria transmission can be described by birth-death processes according to a Moran model with selection. The invasion probability of a low frequency variant is determined by its fitness advantage relative to that of others (*84*).

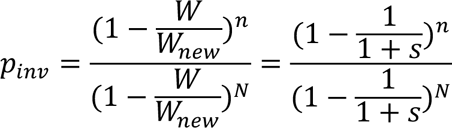

where n denotes the number of copies of a new gene, N, the parasite population size, *W_new_* and W, respectively the fitness of the low-frequency gene versus that of other genes circulating in the population. Conventionally, the fitness of other genes is normalized to be 1, and that of the new gene is denoted as 1+s. We assume that there is a large enough number of alleles so that any mutation or ectopic recombination would lead to a different allele, so that the probability of back mutation to the original allele would be low enough to be negligible. Hence, new variants, generated via mutation or ectopic recombination events, always start with a single copy and n = 1. We consider parasite populations in high-transmission endemic regions, hence *N* ≫ 1. The above equation simplifies to:

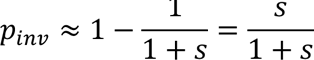

As described in (*85*), in the context of malaria transmission, the fitness of a gene is essentially given by its reproductive number, i.e., the number of offspring genes it produces, R. The reproductive number depends on the transmission rate (b), the transmissibility of the given gene (T), and the typical infection duration (τ) of parasites that carry the given gene.

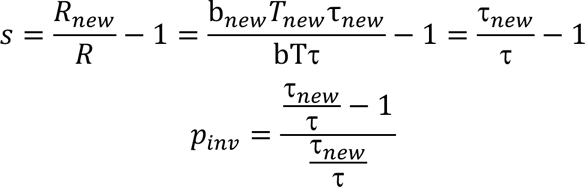

We consider the infection duration of a strain constituted by “average” genes versus the infection duration of a strain constituted by 1 new gene and g-1 average genes where g is the genome size (i.e., the number of *var* genes per individual parasite or repertoire). An average gene is a conceptual entity, and the proportion of susceptible hosts to an average gene (*Ŝ*) is the average of the proportion of susceptible hosts to all circulating genes. Because transmission success of a new gene depends on the total infection duration of the strain that carries it, its invasion probability depends on the overall ensemble of genes. We write here the expected invasion probability for a new gene across the different genome backgrounds it may arise in, by considering that this background consists of average genes. Since the proportion of susceptible hosts to any new gene is 1, we have the following:

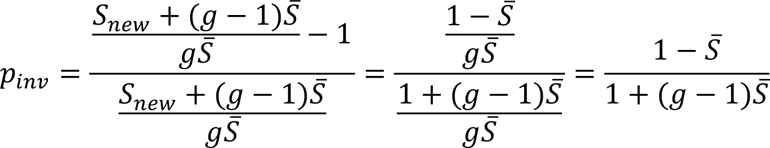

Note that the expression of *p_inv_* derived here is slightly different from, but more rigorous than, the one in (*85*). The two expressions are close under certain conditions, for example, when g is large.

The values for *Ŝ* typical of high-transmission endemic regions are 0.3-0.4 for the low end and around 0.65-0.8 for the high end, based on our ABM simulation outputs. We can use these ranges of *Ŝ* to calculate the invasion probability of a new gene in an individual local parasite population, which together with the innovation rate (primarily through ectopic recombination) and the number of individual local parasite populations (a few for “medium” pools and more than 10 for “large” pools) gives the rate at which common genes are substituted in a regional pool with its specific configuration.

For details on the parameterization of the ABM, see supplementary Materials and Methods (the section “Selection of values for other parameters”) and Data file S1.

### Sampling schemes and measurement error of empirical data

Empirical surveys are carried out at a specific sampling depth and under sparse sampling schemes. When sampling individual hosts and their *var* types in our simulations, we follow the sampling depth implemented in the empirical surveys from Bongo District, namely around 15% of the total human population size (*21*). We further set the sampling period to be once a year for both constant and seasonal transmission (with the former occurring at the end of the year and the latter occurring at the end of the wet/high-transmission season), to emulate the field sampling of hosts primarily at that time of the year, although a few additional surveys were also conducted at the end of the dry (low transmission) season as a baseline for comparison.

The collection of empirical data includes a measurement error, specifically concerning the distribution of the number of non-upsA DBLα sequenced and typed per monoclonal infection, accounting for certain potential sampling errors or imperfect detection of *var* genes (*48, 21*). We incorporate this measurement error when sampling from the simulation output by sub-sampling the number of *var* genes per strain based on this empirical distribution.

Moreover, only individuals with microscopy-positive *P. falciparum* infections (i.e., isolate) were included for sequencing of *var* genes in the empirical surveys from Bongo District. A subset of the longitudinal surveys from Bongo District includes in addition the submicroscopic infections detected by the more sensitive method of PCR (the *18S rRNA* PCR), which resulted in a considerably higher fraction (*35*). Using surveys for which both microscopy and PCR detection were used, we estimated a conversion factor between the proportion of hosts that are microscopy-positive and those that are PCR-positive of 57%. To address the robustness of our results on molecular indicators to such missing data, we repeated the analysis for 57% of all infected hosts originally sampled at the 15% depth.

### Molecular indicators

A 96% DNA sequence identity cutoff was used to define different *var* gene types (*38*). Sequence reads from either the same or different isolates with ≤ 4% sequence differences were grouped into a single consensus or a single type (*21*). We consider the following quantities computed on the basis of *var* gene types and evaluate how these relate to the rebound dynamics of prevalence across the series of simulations for the sustained interventions.

#### Pairwise type sharing (PTS)

The pairwise type sharing (PTS) index describes the degree of shared DBLα types between any two isolates, and is analogous to the Jaccard and Sørensen indices in Ecology (*38, 41, 42*). We use a directional version of this statistic (*29*) defined as 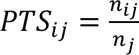 and 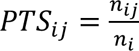 where *n_ij_* is the total number of types shared by isolate i and isolate j, and *n_i_* and *n_j_* are the number of unique types in isolate i and j respectively. This directionality in the formula means that the sharing of a given number of types is relative to the length of the isolate under consideration. A PTS score ranges between 0 and 1, where a score of 0 signifies no shared DBLα types, and a score of 1 indicates complete overlap, between the two isolates. We consider several representative quantiles of the PTS distribution (0.01, 0.31, 0.6, and 0.9) to examine the similarity of isolates that are circulating in the population at the same time or across different times. For comparison of PTS distributions across age groups, we limit the calculation to within the same MOI groups and then aggregate across all MOI values to obtain the final distribution. In other words, within each age group, we compute the PTS between individuals of MOI equal to 1, 2, and so on, and then pool the resulting PTS values together to obtain the final PTS distribution. We do so to minimize the effect of different MOI values on PTS.

#### Multiplicity of infection (MOI)

Because *Plasmodium* parasites reproduce asexually as haploid stages within human hosts, signatures of polymorphic genotypes are evidence of multiclonal infection. While any polymorphic marker should thus be suitable in theory for estimating MOI, the typically high number of multiclonal infections in high-transmission endemic regions limits our ability to accurately do so in practice. This is mainly because the diversity of the polymorphic markers can be limited, with two parasites sharing the same marker but carrying distinct sets of antigen-encoding genes. The *var* multigene family provides one solution. Because of the large number of different genes in local populations and the effect of immune selection (negative frequency-dependent selection), repertoires of *var* genes in individual genomes exhibit very limited overlap (*23, 29*). As a result, a constant repertoire size or number of non-upsA DBLα types in a parasite genome can be used to convert the number of types sequenced in an isolate to an estimate of MOI (*24, 35*). In its simplest form of considering a typical repertoire length per parasite, the approach neglects the measurement error introduced by targeted PCR and amplicon sequencing of *var* genes in an infection. Recently, the method was extended to a Bayesian formulation that does consider this error and provides a posterior distribution of different MOI values for each sampled individual (*21*). We have previously documented the major steps of the Bayesian approach, compared two ways of obtaining a MOI distribution at the population level (by either pooling the maximum a posteriori MOI estimates or calculating a mixture distribution) (*21*), and examined the impact of different priors on the final MOI distribution at the population level. Here, we obtain the estimated MOI distribution at the population level from pooling the maximum a posteriori MOI estimates and from using a uniform prior for individuals.

Due to the commonness of multi-genomic infections in high-transmission regions, host prevalence levels are not a good indicator of parasite population size. Summing MOI estimates from this improved “*var*coding” approach over sampled individuals with infections, enables the calculation of census population size for the parasite or the total number of infections of relevance to transmission events (*21*).

## List of Supplementary Materials

Present a list of the Supplementary Materials in the following format.

Materials and Methods

Fig. S1 to S22 for multiple supplementary figures

Data file S1 (Excel files)

References (*70-89*) (numbers for references only cited in SM)

## Supporting information

DataFileS1

## Data Availability

The sequences utilized in this study are publicly available in GenBank under BioProject Number: PRJNA 396962. All additional data associated with this study are available in the main text, the supplementary information, or upon reasonable request to the authors. For information on the simulation code and analysis scripts, please see https://github.com/qzhan321/Intervention.

## Acknowledgments

We wish to thank the participants, communities, and the Ghana Health Service in Bongo District, Ghana for their willingness to participate in the study of empirical data. We would like to thank the field teams in Bongo for their technical assistance in the field, as well as the laboratory personnel at the Navrongo Health Research Centre for their expertise and for undertaking the sample collections and parasitological assessments.

## Author contributions

Conceptualization: QZ, MP, KPD

Methodology: QZ, MP

Investigation: QZ

Visualization: QZ

Funding acquisition: MP, KPD

Project administration:

Supervision: MP, KPD

Writing – original draft: QZ

Writing – review & editing: QZ, MP, KD, KT, QH

## Supplementary Material

We describe details of key assumptions and processes of the ABM, including extensions implemented to the model for this work.

### *Var* genes and repertoire structure

Each parasite genome consists of a specific combination of 45 non-upsA *var* genes in the simulation. The size of the repertoire is based on the median number of non-upsA DBLα types identified in the previously reported 3D7 laboratory data (*35, 69*). The grouping of *var* genes (i.e., upsA and non-upsA (upsB and upsC)) is based on their structural information (i.e., the semi-conserved upstream promoter sequences or ups) (*59, 60, 61*). Each empirically sampled parasite carries both types of *var* genes in a fairly constant proportion, with the majority (∼80%) being non-upsA groups (*21, 22, 24, 55, 59*). Although there is evidence indicating functional differences between the two groups, the groupings do not necessarily correlate with function. Genes of the same group can differ in their functional properties (*57, 59*). Genes of different groups may both be significantly associated with a particular type of infection symptom (*58, 59*). Within-group functional heterogeneity and variation, and cross-group functional similarity exist. In addition, the mechanism underlying the fairly constant proportion of two structural groups in empirical samples across time and geographical locations remains to be properly understood. It is therefore unclear how to model functional divergence of *var* groups in a rigorous and parsimonious way, and whether assumptions made on the particular coarse-graining apply to the population dynamics of the disease. We consider here only non-upsA types in our simulation and discuss extensions introducing the more conserved upsA types. The non-upsA DBLα sequences are in general ∼20 times more diverse and less conserved among repertoires than the upsA DBLα sequences (*21*), making them suitable and sufficient for MOI estimation based on deep sampling of sequences of *var* genes, as described in the section “Molecular indicators” of Materials and Methods.

### Repertoire transmission

Mosquito vectors are not explicitly represented as agents in the model. Instead, we consider an effective contact rate (hereafter, the transmission rate, which determines the times of local transmission events (exponentially distributed according to a Poisson process). At these times, a donor and a recipient host are selected randomly, and successful transmission occurs only if the donor carries active blood-stage infections, and the recipient has not reached a carrying capacity of the liver stage of infection (see the following section “Within-host dynamics”). Co-infection reduces transmission success of each individual parasite strain. When the donor is infected with multiple strains in the blood stage, then the transmission probability of each strain is reduced by a factor equal to the number of co-infecting strains (*70, 72, 73*). We apply a delay between a successful transmission event and the transmitted strains in the recipient host starting to express their *var* genes and becoming infectious, to account for stages of the life cycle in the vector and in the human host that are not modeled explicitly (see the following section “Within-host dynamics”).

Note that for simplicity, when a transmission event occurs, our model considers that all infectious “bites” (i.e., for a donor host that carries active blood-stage infections and a recipient host that has liver capacity), result in infection of the recipient host. The probability of infection given infectious bites can be considerably less than 1 in nature. For example, a bite with an adequate volume of sporozoites has been shown to have a probability of infecting human hosts of around 10% under certain constraints (*74*). In general, when transmission is high and multi-genomic infections are common, this probability can be even much lower than 10% (75). Because for our dynamics, this parameter represents a rescaling, it does not need to be incorporated explicitly, as long as for comparison of entomological inoculation rates (the number of infectious bites received by an individual over a given time period or EIR) in our model and field estimates, the former is divided by the transmissibility probability. Additionally, our model generates values of the force of infection (the number of new infections acquired by an individual host over a given time interval or FOI). For pre-intervention dynamics, annual FOI values are within the range of 9-15, directly comparable to the measures and estimates from the field for high-transmission endemic regions in sub-Saharan Africa (*75*).

### Ectopic recombination during the asexual blood stage of infection

Ectopic recombination is a major mechanism of *var* genes’ diversification, occurring during both the sexual and asexual stages. For simplicity, we only model ectopic recombination among genes within the same genome during the asexual stage as follows. Two genes are first selected randomly from a given strain. Then, the location of the recombination breakpoint is randomly chosen. Under normal recombination, both genes have their alleles swapped and there is a certain probability that new alleles are created in the process. Under conversion, the second gene remains unchanged. In the current implementation, we assume all ectopic recombination events result in normal recombination rather than gene conversion. Newly recombined genes have a probability of being functional (i.e., viable), dependent on the similarity of their parental genes and the locations of the breakpoints (*76*), and they will replace their parental ones. Non-functional genes will not express and get deactivated immediately and hence do not increase infection duration.

### Meiotic recombination (or outcrossing) during the sexual stage of infection

Meiotic recombination occurs between strains during sexual replication inside the mosquito vector. When multiple strains co-infect vectors, they can either remain unchanged or recombine with each other. Although the association of physical locations and major groups of *var* genes is established, orthologous gene pairs between two strains are often unknown. Therefore, we implement recombination between strains as a process in which genes are randomly selected out of all the original genes from the two strains pooled together. As physical locations of *var* genes can be mobile through ectopic recombination and gene conversions, this assumption is a reasonable simplification of the meiotic recombination process. The resulting offspring strains share some fraction of their *var* genes with the two parental strains. Outcrossing creates relatedness and increases similarity between parental and offspring strains.

### Within-host dynamics

Each strain is individually tracked through its entire life cycle, encompassing the liver stage and asexual blood stage in the human host, and the sexual stage in the mosquito. As we do not explicitly model mosquitoes, we delay the expression of each strain in the recipient host by 14 days to account for the time required for the sexual and liver stages. Specifically, the infection of the host is delayed 7 days to account for the time required for gametocytes to develop into sporozoites within mosquitoes. When a host is infected, the parasite remains in the liver stage for an additional 7 days before being released as merozoites into the bloodstream, invading red blood cells and starting the expression of the *var* repertoire (*77*). The expression of genes in the repertoire is sequential and the infection ends when the whole repertoire is depleted. During repertoire expression, the host is considered infectious with the active strain. Only these active “blood-stage” strains are transmissible to another host. The deactivation rates of the genes are controlled by host specific immunity. When one gene is actively expressed, host immunity “checks” whether it has seen any of the two epitopes in its infection history. If both epitopes have been previously encountered, the gene gets deactivated immediately. The duration of the active period of a gene is thus proportional to the number of unseen epitopes of the gene. After the gene is deactivated, the host adds the deactivated two epitopes to its immunity memory. The next new gene from the repertoire then becomes immediately active and the strain is cleared from the host when the whole repertoire of *var* genes is depleted. The total duration of infection of a particular repertoire is therefore proportional to the number of unseen epitopes by the infected host aggregated across all individual *var* genes of that repertoire. The immunity toward a certain epitope wanes at a certain rate.

In addition, we consider a carrying capacity for both the liver- and blood-stage infection respectively. We adopt 20 for both quantities because this has been the maximum MOI observed in the empirical data from Bongo (*21*). When the number of strains in the liver stage exceeds the specified carrying capacity, the host no longer receives additional infections when it is selected as the recipient host for transmission events. When the number of strains in the blood stage exceeds the carrying capacity, strains in the liver stage are not released into the bloodstream and fail to “transition to the blood-stage”. In this implementation, we assume these strains are lost instead of queuing indefinitely (*78*).

As previously described in the section “Repertoire transmission”, we make sure that the chosen values for transmission rates or associated FOI in the simulation are high enough to capture disease transmission in high-transmission endemic regions but still sufficiently low to preclude significant void bites (because of exceeding carrying capacities).

### Selection of values for other parameters

#### Immunity loss rate

Values of this parameter can be obtained from the data on the treatment of neurosyphilis patients with malaria infections in the 1950s to 1960s (87, 88). Their antibody titers were tracked through observation periods of up to decades following the termination of their very first inoculation. During these periods, these individuals usually did not receive repeated, either homologous or heterologous, infections, which can boost immunity and confound the immunity loss rate. In (*86*), the persistence of the antibody response against malaria was studied. Sera from 95 patients having had induced infections with *P. falciparum*, *P. malariae*, *P. vivax,* or *P. ovale* were tested for presence of specific antibody using the indirect fluorescent antibody methods. The intervals between termination of the infections and acquisition of the sera ranged from approximately 6 months to 26 years. Among these patients, 14 of them had a past history of *P. falciparum* infection. A specific serum dilution high enough to be considered positive was found as late as 8 years after the termination of *P. falciparum* infection. In general, the median response decreased sharply when three years had elapsed from the termination of infections.

In (*87*), a different set of 7 patients was followed to track persistence of the antibody response only up to 20 months post inoculation. During the 8-month period (between 12 months and 20 months post inoculation), 3 of the titers remained unchanged, 2 decreased by one dilution, 1 decreased by two dilutions, and 1 decreased by three dilutions. Roughly titers decrease by 1 dilution for every 8 months.

Combining information from both studies, we set the mean immunity duration to be 3 years in the simulation.

#### Number of epitopes (alleles) per locus

(*54*) presented a new method that identifies nine highly variable regions (HVRs) and studied 307 amino acid sequences from the DBLα domain of the *var* genes of seven *P. falciparum* isolates. Two HVRs (5 and 6) correspond to the DBLα tag sequences that were amplified in our field isolates from Bongo District. For each HVR, the study constructed a network in which each sequence is a node, and two nodes are linked if they share an exact match of significant length of the HVR. Using a community partition inference algorithm, the authors found that the number of components ranges from 1 to 106 across different HVRs, with a mean around 30. Specifically, 10 and 8 components were found for HVRs 5 and 6. We therefore assume a reduction of an order of 10 between the number of genes and the number of alleles (epitopes) at each of the two loci.

#### Ectopic recombination rate

In (*26*), the average total number of *var* recombination events per life cycle (2 days) was estimated as 2 × 10^−3,^. Given that there are around 60 *var* genes per parasite genome and hence 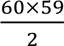 different combinations of *var* gene pairs for a recombination event to happen, the previous value amounts to about 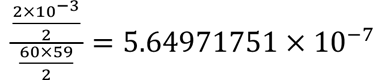 recombination events per *var* gene pair per day. Note that there was some heterogeneity and hierarchy observed in the study so that certain pairs of *var* genes from a specific structural group recombined more frequently than others and recombination events are more likely to be observed between genes from the same structural group than the ones from different structural groups. We do not consider such a variation across all gene pairs but use the above-calculated mean value in our simulation output.

Ectopic recombination events do not necessarily generate new alleles, and they happen more frequently between similar homology blocks, the exchange of which is less likely to result in antigenically new offspring alleles. We test a high and a low probability of generating antigenically new alleles per ectopic recombination event, i.e., 0.5 (the upper bound) and 0.16, in other words, 1 in 2 versus 1 in 6 ectopic recombination events. The results and patterns are robust across the two probability values. We thus report the results and patterns based on simulation output which assume that a 0.16 probability (1 in 6 ectopic recombination events) will result in a new allele.

#### Mutation rate

The same study (*26*) estimated that a weighted average mutation rate of 4.07 × 10^−10^, 3.63 × 10^−10^, 3.78 × 10^−10^ SNPs per erythrocytic life cycle per nucleotide for 3D7, Dd2, and HB3 respectively (*26*). Based on (*88*), we assume 5AA mutations will result in a new type. Each allele can represent either one HVR (25-57 bp) or half of the exon 1 of *var* genes (around 2500 bp). The average mutation rate per allele per day is of the order of 10^−9^ to 10^−7^. We assume it to be 10^−8^in our simulation.

---

**Fig. S1.**
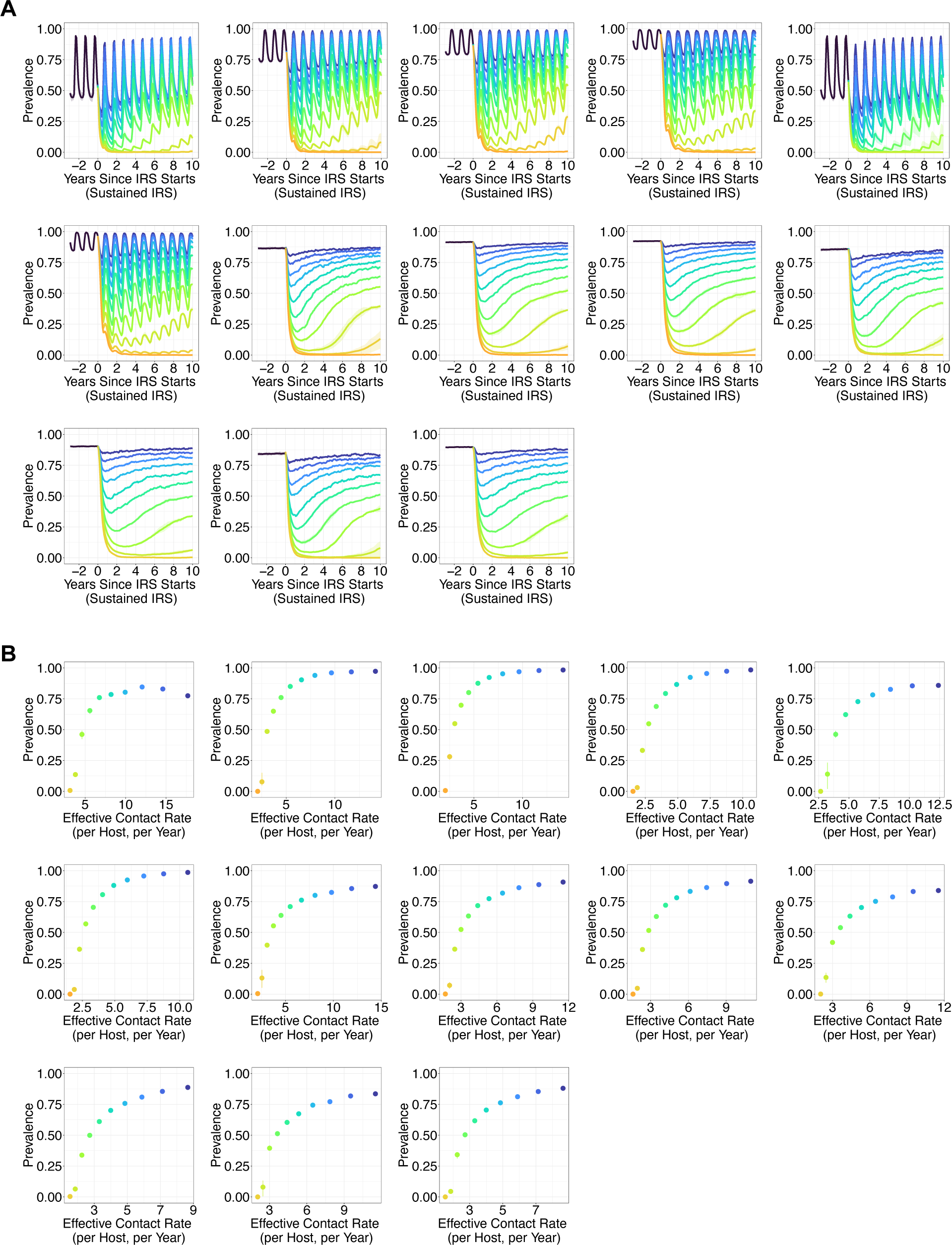
(**A**) Rebound dynamics of prevalence across time under a series of sustained IRS interventions, for closed, semi-open, and regionally-open systems simulated with additional combinations of parameter values than the one shown in Fig. 2A and B. The dynamics can be categorized into three regimes: the fast-rebound, the transition, and the slow-rebound regime. The top row from left to right are seasonal and closed systems (scenario I in Fig. 1B), seasonal and semi-open systems with a baseline migration rate (scenario III), seasonal and semi-open systems with a high migration rate (an order higher than the baseline, scenario IV), seasonal and regionally-open systems with a baseline migration rate and a large pool (scenario VIII), seasonal and regionally-open systems with a high migration rate and a medium size pool (scenario IX). The middle row from left to right are seasonal and regionally-open systems with a high migration rate and a large pool (scenario X), non-seasonal and closed systems (scenario II), non-seasonal and semi-open systems with a baseline migration rate (scenario V), non-seasonal and semi-open systems with a high migration rate (scenario VI), non-seasonal and regionally-open systems with a baseline migration rate and a medium size pool (scenario XI). The bottom row from left to right: non-seasonal and regionally-open systems with a baseline migration rate and a large pool (scenario XII), non-seasonal and regionally-open systems with a high migration rate and a medium pool (scenario XIII), non-seasonal and regionally-open systems with a high migration rate and a large pool (scenario XIV). (**B**) Long-term rebound prevalence levels against transmission rates during sustained IRS interventions. Specifically, we plot prevalence levels at the 10th year into the sustained IRS intervention, sampled either at the end of wet/high-transmission season for seasonal transmission, or at the end of the year for constant transmission. Different colors indicate varying levels of intervention intensity.

**Fig. S2.**
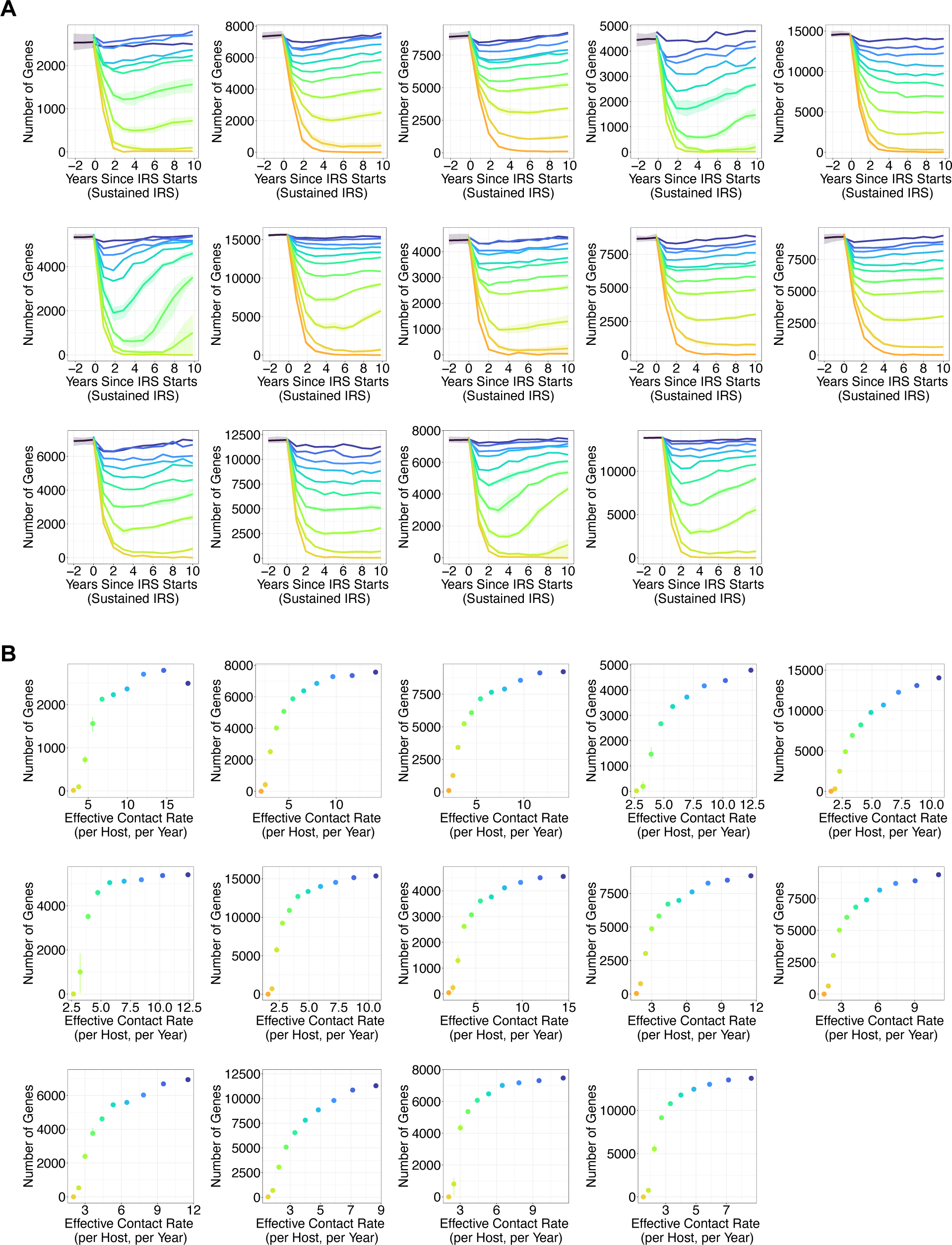
(A) Rebound dynamics of *var* gene diversity measured as richness across time under a series of sustained IRS interventions. The top row from left to right are seasonal and closed systems (scenario I in Fig. 1B), seasonal and semi-open systems with a baseline migration rate (scenario III), seasonal and semi-open systems with a high migration rate (an order higher than the baseline, scenario IV), seasonal and regionally-open systems with a baseline migration rate and a medium size pool (scenario VII), seasonal and regionally-open systems with a baseline migration rate and a large pool (scenario VIII). The middle row from left to right are seasonal and regionally-open systems with a high migration rate and a medium size pool (scenario IX), seasonal and regionally-open systems with a high migration rate and a large pool (scenario X), non-seasonal and closed systems (scenario II), non-seasonal and semi-open systems with a baseline migration rate (scenario V), non-seasonal and semi-open systems with a high migration rate (scenario VI). The bottom row from left to right: non-seasonal and regionally-open systems with a baseline migration rate and a medium size pool (scenario XI), non-seasonal and regionally-open systems with a baseline migration rate and a large pool (scenario XII), non-seasonal and regionally-open systems with a high migration rate and a medium pool (scenario XIII), non-seasonal and regionally-open systems with a high migration rate and a large pool (scenario XIV). (B) Long-term rebound dynamics of richness against transmission rates. Specifically, we plot richness levels at the 10th year into the sustained IRS intervention, sampled either at the end of wet/high-transmission season for seasonal transmission, or at the end of the year for constant transmission. For semi-open systems, we plot the total richness of the two explicitly coupled local parasite populations as we sample both populations. For closed and regionally-open systems, we plot the richness of the focal parasite population only.

**Fig. S3.**
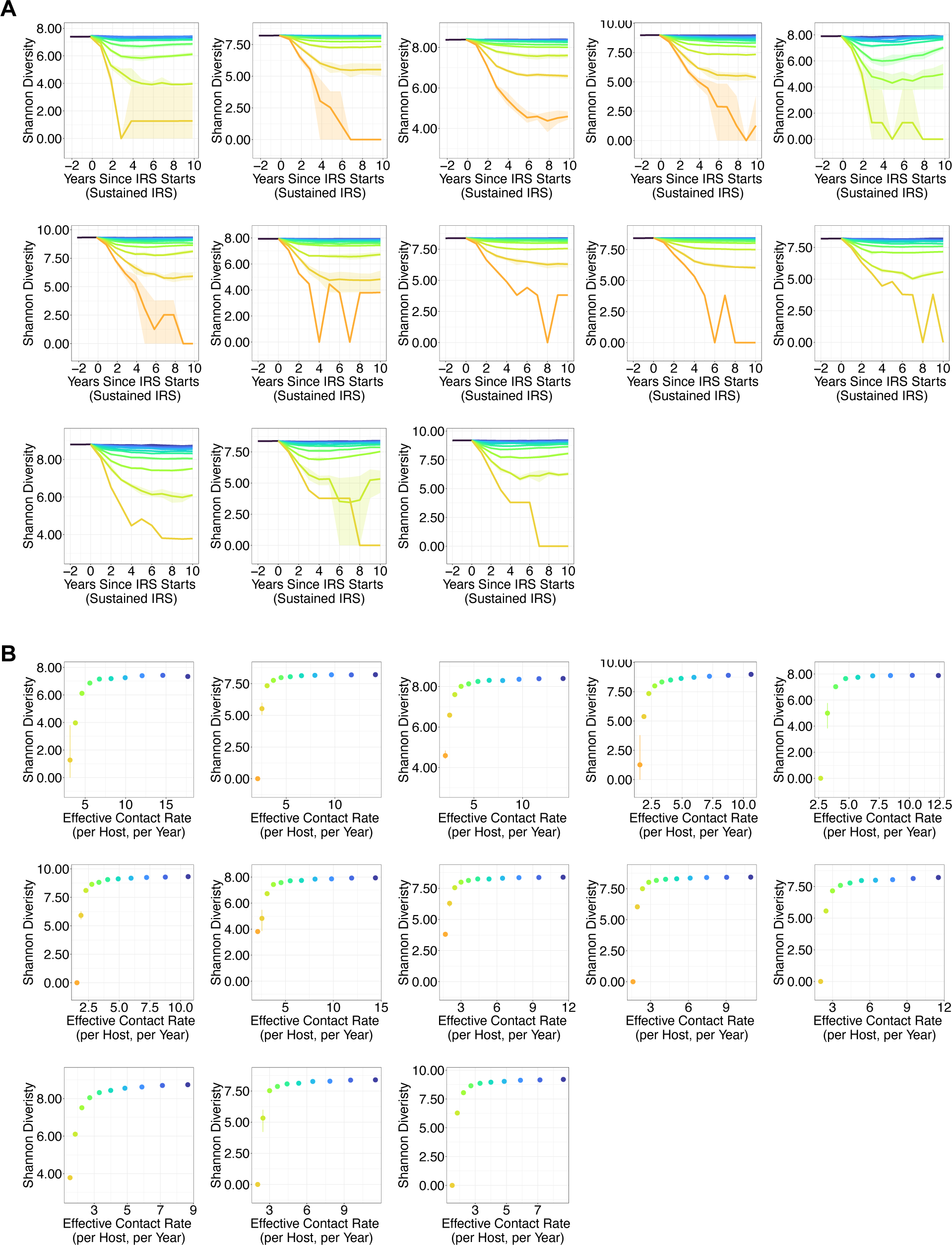
(A) Rebound dynamics of *var* gene diversity measured by the Shannon Diversity Index across time under a series of sustained IRS interventions, for closed, semi-open, and regionally-open systems simulated with additional combinations of parameter values than the one shown in Fig. 2C and 2D. The top row from left to right are seasonal and closed systems (scenario I in Fig. 1B), seasonal and semi-open systems with a baseline migration rate (scenario III), seasonal and semi-open systems with a high migration rate (an order higher than the baseline, scenario IV), seasonal and regionally-open systems with a baseline migration rate and a large pool (scenario VIII), seasonal and regionally-open systems with a high migration rate and a medium size pool (scenario IX). The middle row from left to right are seasonal and regionally-open systems with a high migration rate and a large pool (scenario X), non-seasonal and closed systems (scenario II), non-seasonal and semi-open systems with a baseline migration rate (scenario V), non-seasonal and semi-open systems with a high migration rate (scenario VI), non-seasonal and regionally-open systems with a baseline migration rate and a medium size pool (scenario XI). The bottom row from left to right: non-seasonal and regionally-open systems with a baseline migration rate and a large pool (scenario XII), non-seasonal and regionally-open systems with a high migration rate and a medium pool (scenario XIII), non-seasonal and regionally-open systems with a high migration rate and a large pool (scenario XIV). (B) Long-term rebound dynamics of the Shannon Diversity Index against transmission rates. Specifically, we plot diversity levels at the 10th year into the sustained IRS intervention, sampled either at the end of wet/high-transmission season for seasonal transmission, or at the end of the year for constant transmission. For semi-open systems, we plot the Shannon Diversity Index of the two explicitly coupled local parasite populations as we sample both populations. For closed and regionally-open systems, we plot the Shannon Diversity Index of the focal parasite population only. Prevalence in figs. S1, *var* gene diversity measured as richness in figs. S2, and *var* gene diversity measured by the Shannon Diversity Index here all demonstrate a non-linear behavior as a function of the transmission rates. The fast-rebound regime occurs for a wide range of transmission rates during intervention, whereas the transition and slow-rebound regimes occur for a narrow region along the gradient of transmission rates.

**Fig. S4.**
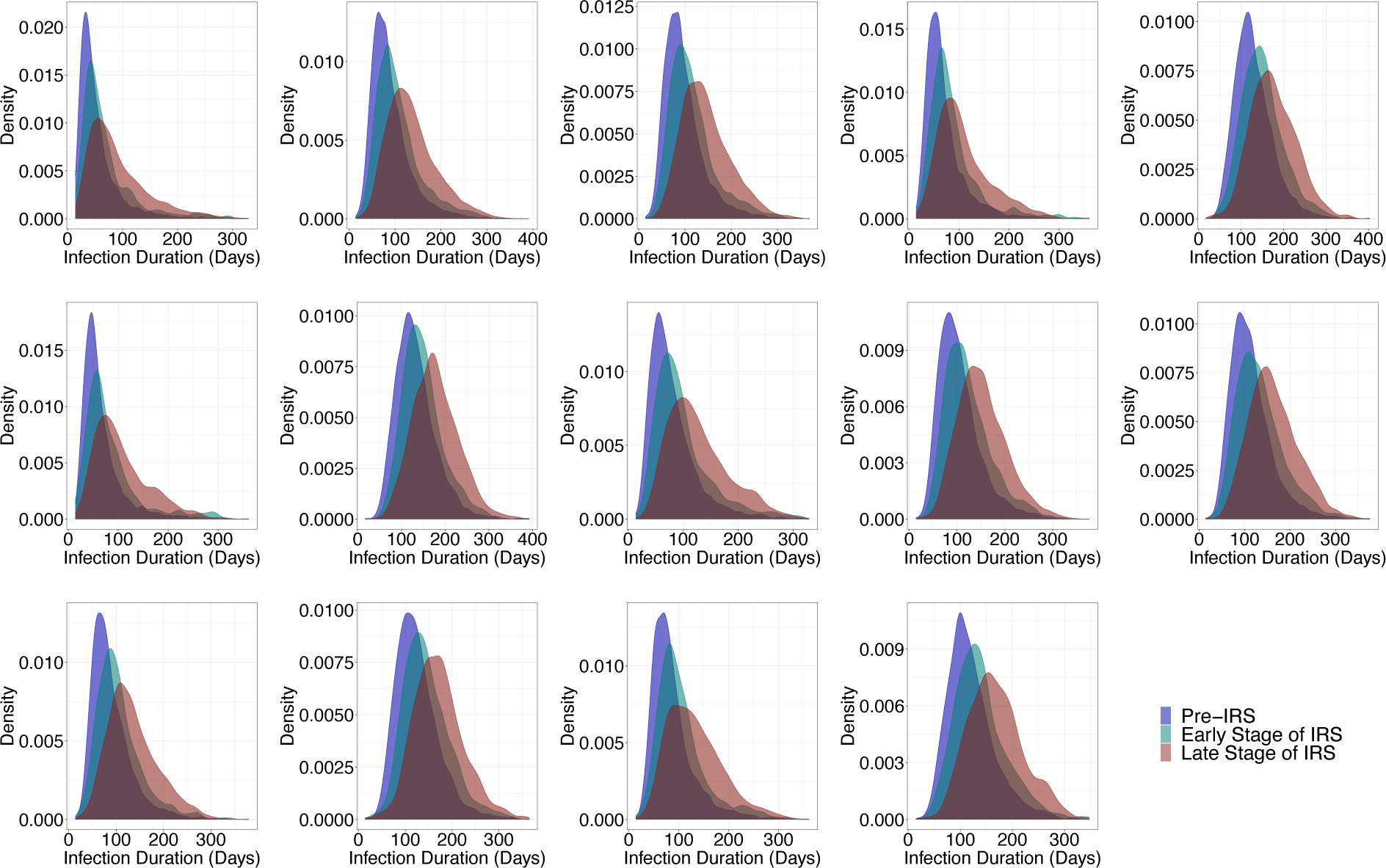
The shift in distributions of infection duration from before to immediately following to towards the end of a particular sustained IRS intervention. For illustration purposes, we present here I-4 (for its level of transmission intensity, see Fig. 1D). Specifically, we consider infections which start and end within three years before intervention, those which start and end within three years after intervention is applied, and those which start and end within the last three years of the decade-long intervention phase. The distribution keeps shifting to the right towards longer values throughout the intervention phase relative to that before intervention. This pattern also holds for intervention runs of different coverages than I-4 shown here. The degree of the shift increases as the coverage of the intervention increases. Infections with longer duration are more likely get transmitted than infections with shorter infection duration, due to a reduction in transmission rates. Therefore, the entire infection duration distribution shifts to the right. In addition, the overall shape of the distribution during intervention becomes more heterogeneous and widely spread out. The change in the distribution of infection duration results from a new relationship between hosts’ heterogeneous immunity profiles and systems’ surviving diversity, which can effectively compensate for a reduction in transmission intensity for systems under low- or mid-coverage interventions. These perturbed systems start to rebound early into intervention. However, under very high-coverage interventions, transmission intensity drops to levels that are too low. Consequently, few infections can survive longer than the average waiting time between consecutive bites and transmission events. Reduction in transmission cannot be effectively compensated for. These perturbed systems remain at low prevalence and diversity levels for long. Note that in general duration infection is longer for systems with high circulating diversity, for example, regionally-open systems with a large pool, than systems with lower circulating diversity, for example, closed systems. This is intuitive as high circulating diversity promotes immune evasion of parasites and lengthens infection duration. The top row from left to right are scenario I (Fig. 1B), III, IV, VII, VIII. The middle row from left to right are scenario IX, X, II, V, VI. The bottom row from left to right are scenario XI, XII, XIII, XIV.

**Fig. S5.**
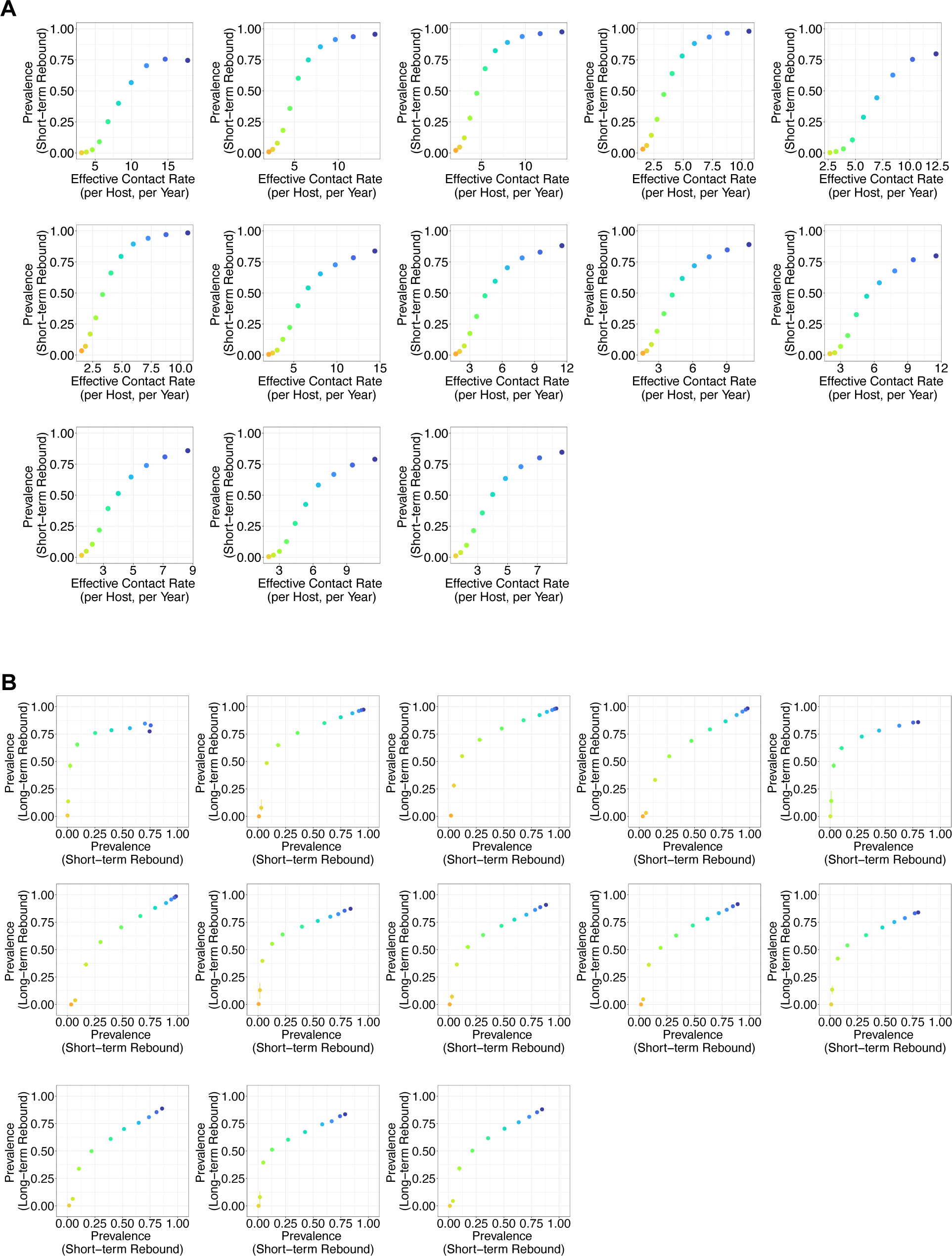
(A) Short-term rebound prevalence levels against transmission rates of the series of sustained intervention. We specifically plot prevalence levels at the 2nd year into the series of sustained intervention (sampled at the end of wet/high-transmission season for seasonal transmission and at the end of the 2nd year for constant transmission). The top row from left to right are scenario I (Fig. 1B), III, IV, VIII, IX. The middle row from left to right are scenario X, II, V, VI, XI. The bottom row from left to right are scenario XII, XIII, XIV. (B) The relationship between short-term and long-term (at the 10th year into intervention) rebound prevalence levels. The relationship between these two types of prevalence levels is non-linear, and the degree of nonlinearity varies across different spatial configurations and parameter values.

**Fig. S6.**
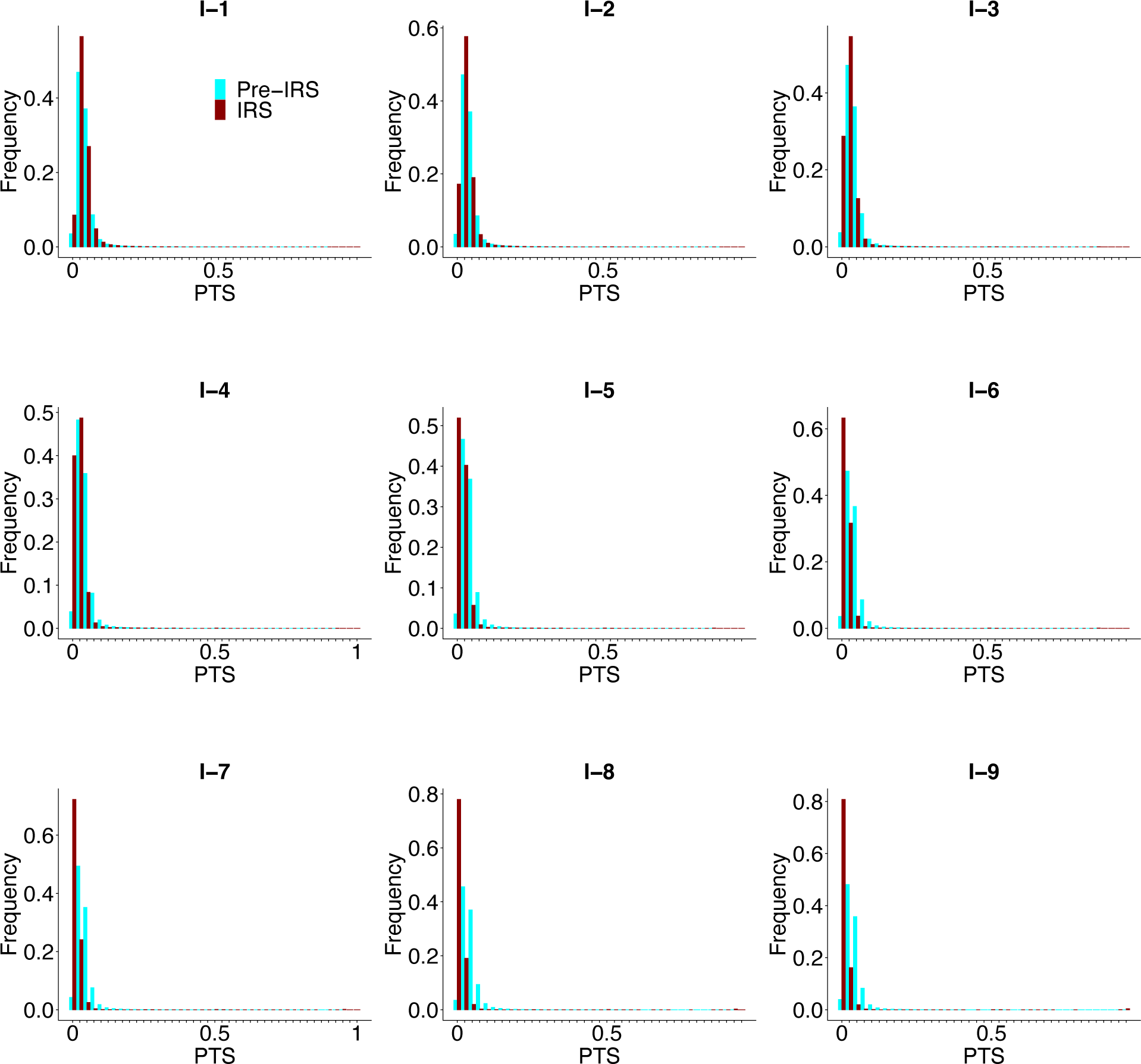
The emergence of the second mode at complete overlap becomes insignificant when intervening systems with high circulating diversity, for example, seasonal and regionally-open ones with a high migration rate and a large regional pool (scenario X in Fig. 1B). Plotted here are PTS values calculated based on true samples, i.e., the ones without sampling limitations (missing data issue and measurement error) representative of those encountered in the collection of field data.

**Fig. S7.**
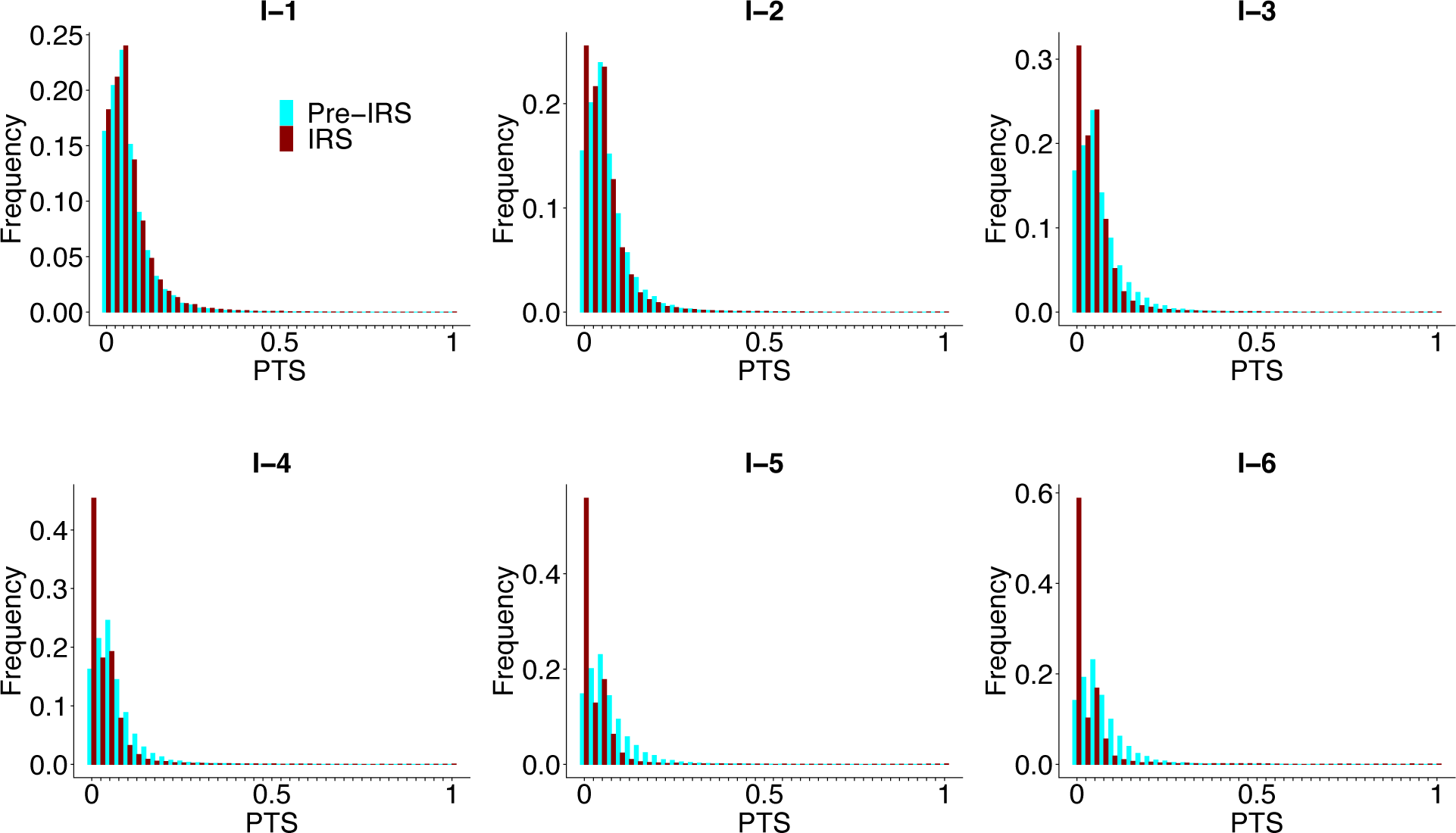
The pairwise type sharing or PTS distribution during sustained intervention for regionally-open systems with a baseline migration rate and a medium pool (scenario VII in Fig. 1B). When sampling the simulation output for PTS calculation, we implemented sampling schemes and sampling limitations representative of those encountered in the collection of field data. The emergence of the second mode disappears (see section “Sampling schemes and measurement error of empirical data” in Materials and Methods). The case without sampling limitations (missing data issue and measurement error) is shown in Fig. 3D.

**Fig. S8.**
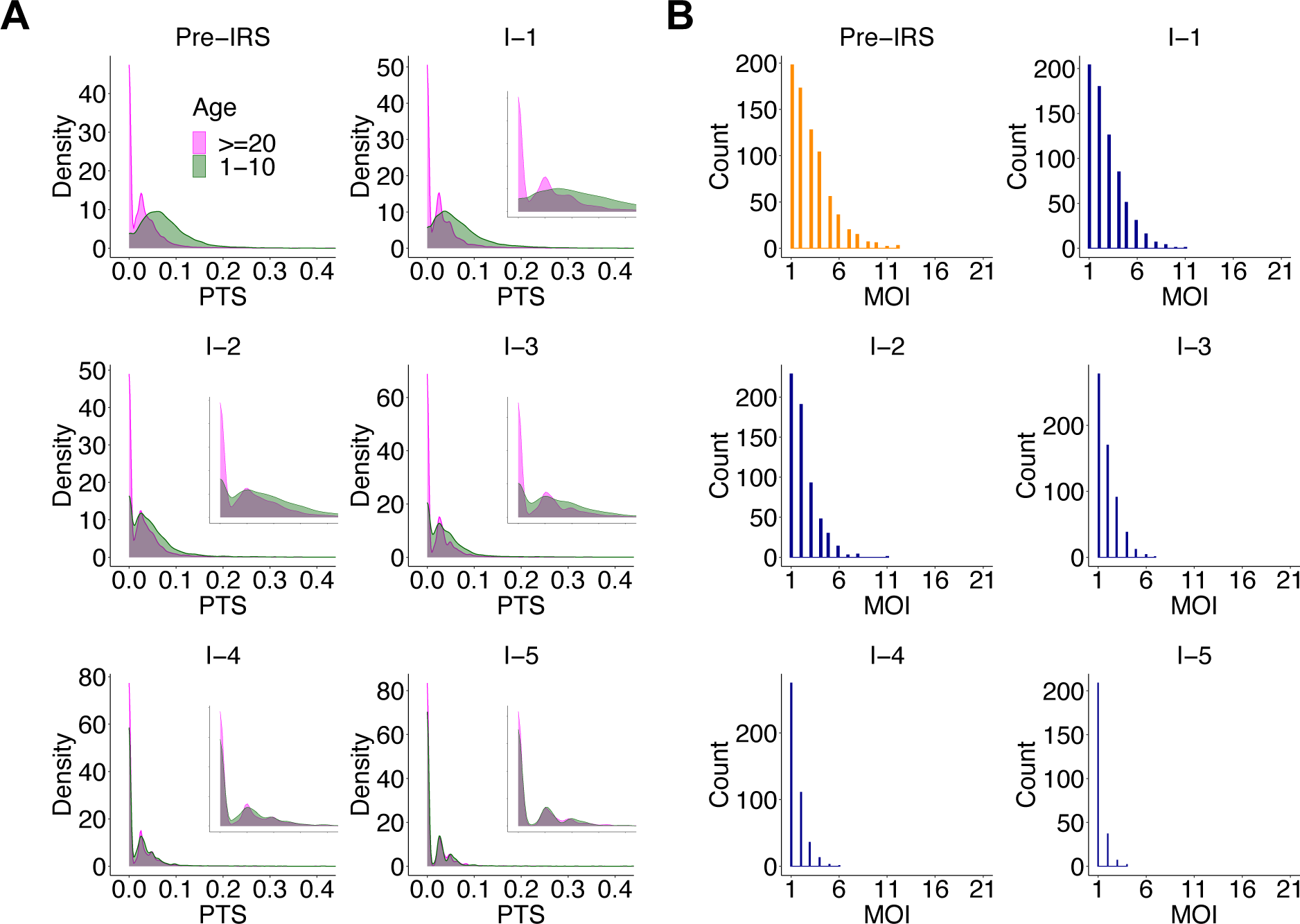
Molecular indicators of the proximity of the perturbed local system to the transition regime for seasonal regionally-open systems with a medium pool and a baseline migration rate (scenario VII in Fig. 1B). (A) As the system approaches the transition regime, the difference between PTS distributions across age groups is significantly reduced and even disappears. In particular, the two distributions almost completely overlap at the lowest mode around 0. The x-axis range for inset figures is 0-0.1. (B) MOI distribution starts to center around 1 with the majority of infections being either mono-clonal or multi-genomics with two genetically distinct parasites. These two molecular indicators are robust under implemented sampling schemes (sampling frequency and depth) and sampling limitations representative of those encountered in the collection of field data (the missing data issue and measurement error). The case without sampling limitations (the missing data issue and measurement error) is shown in Fig. 4.

**Fig. S9.**
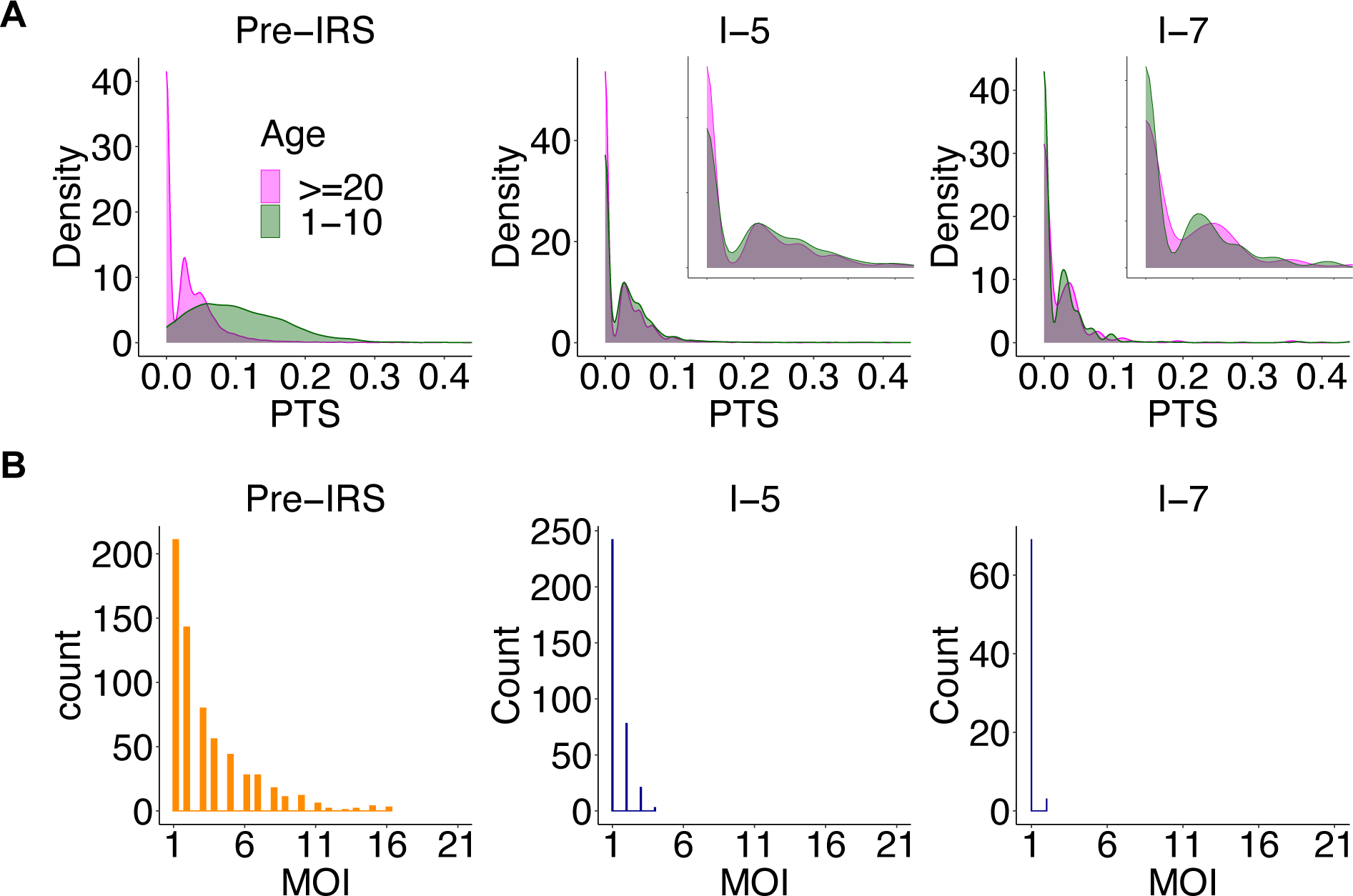
The two molecular indicators for seasonal closed systems (scenario I in Fig. 1B). For illustration purposes, we show the pre-IRS case, an example IRS under which the system approaches the transition regime, and another example IRS under which the system falls into the transition or slow-rebound regime. (A) As the system approaches the transition regime, the difference between PTS distributions across age groups is significantly reduced and even disappears. In particular, the two distributions almost completely overlap at the lowest mode around 0. The x-axis range for inset figures is 0-0.1. (B) MOI distribution starts to center around 1 with the majority of infections being either mono-clonal or multi-genomic with two genetically distinct parasites. These two molecular indicators are robust under implemented sampling schemes (sampling frequency and depth) and sampling limitations representative of those encountered in the collection of field data (the missing data issue and measurement error).

**Fig. S10.**
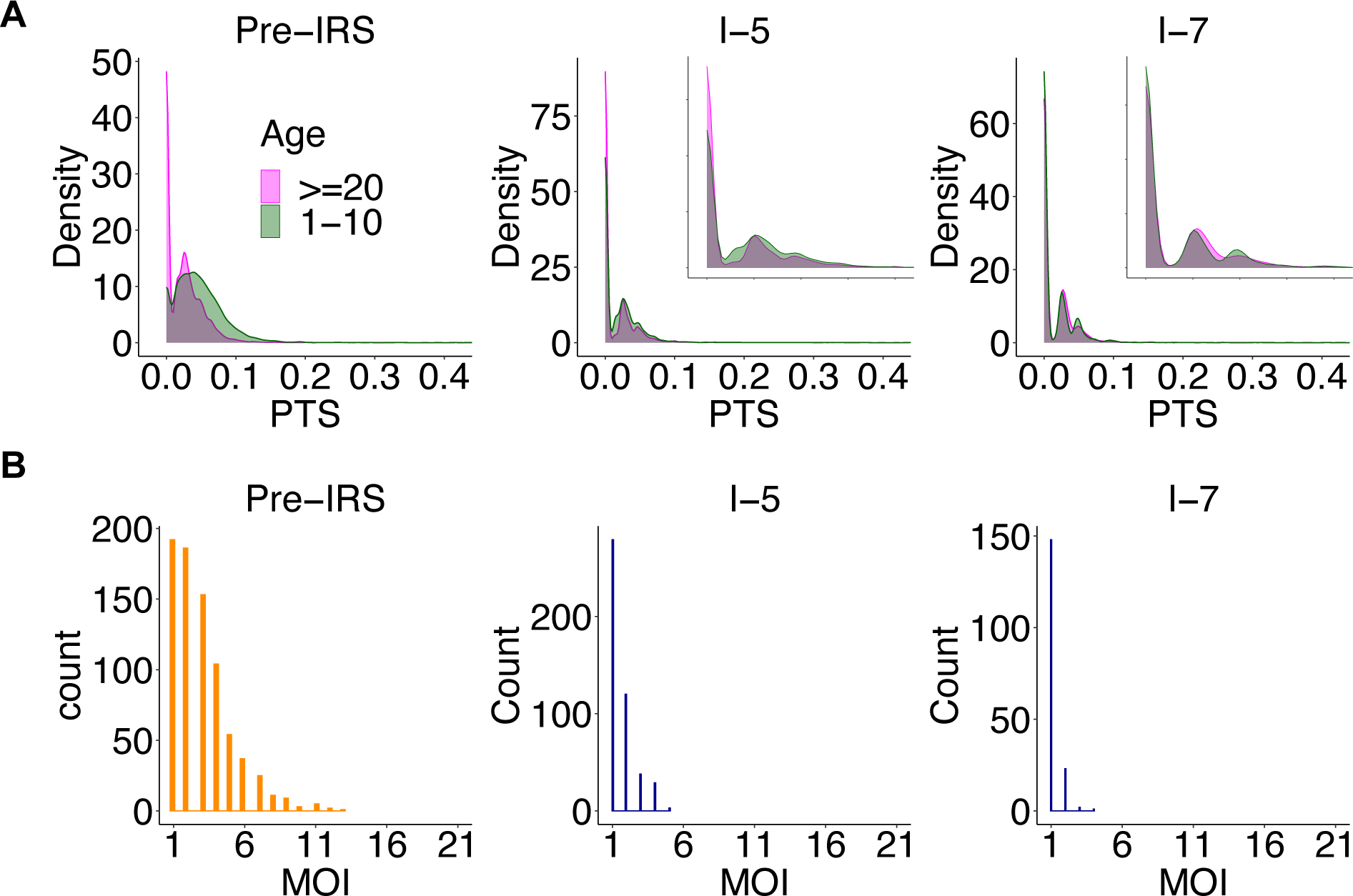
The two molecular indicators for non-seasonal closed systems (scenario II in Fig. 1B). For illustration purposes, we show the pre-IRS case, an example IRS under which the system approaches the transition regime, and another example IRS under which the system falls into the transition or slow-rebound regime. (A) As the system approaches the transition regime, the difference between PTS distributions across age groups is significantly reduced and even disappears. In particular, the two distributions almost completely overlap at the lowest mode around 0. The x-axis range for inset figures is 0-0.1. (B) MOI distribution starts to center around 1 with the majority of infections being either mono-clonal or multi-genomic with two genetically distinct parasites. These two molecular indicators are robust under implemented sampling schemes (sampling frequency and depth) and sampling limitations representative of those encountered in the collection of field data (the missing data issue and measurement error).

**Fig. S11.**
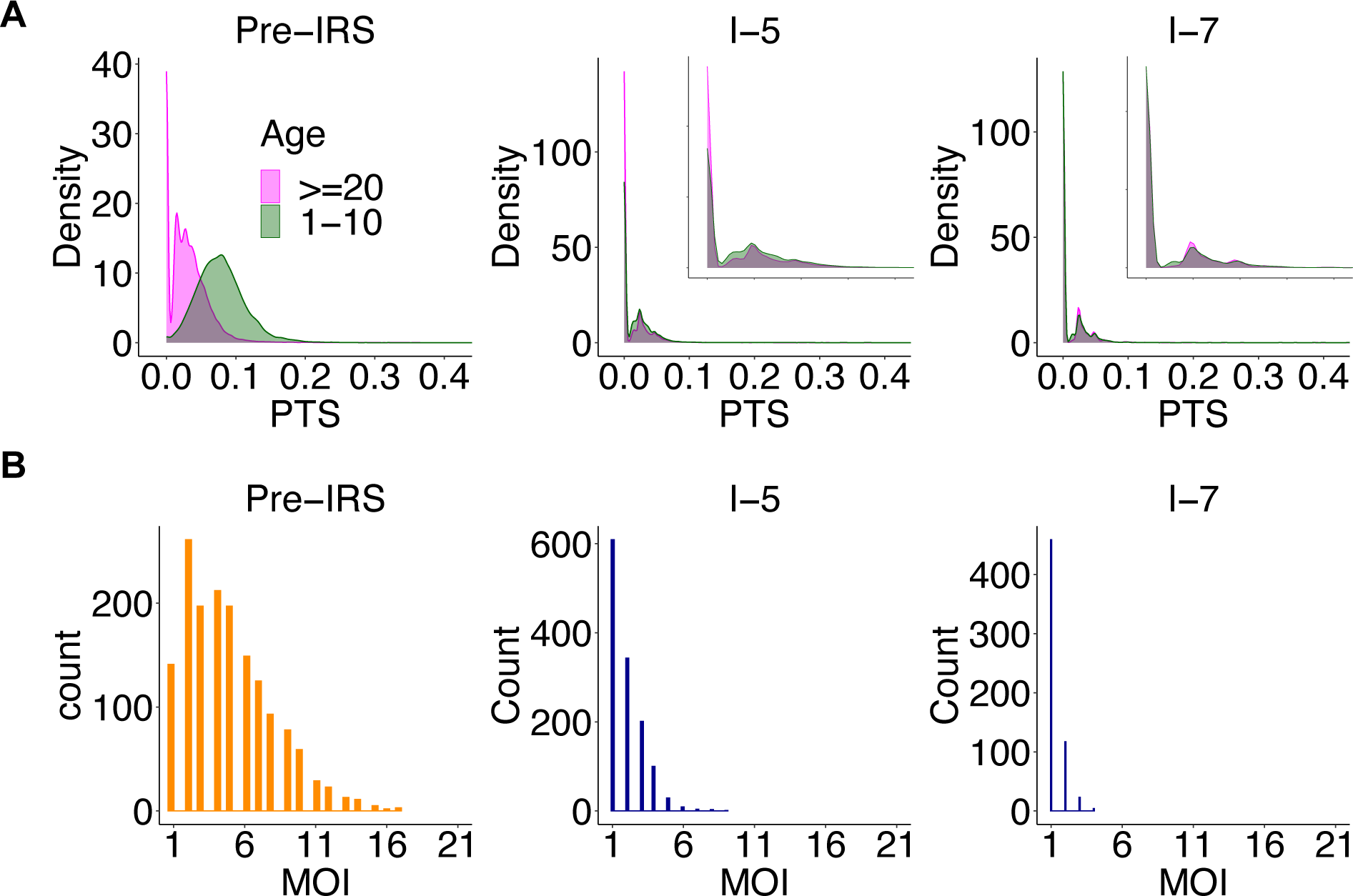
The two molecular indicators for seasonal semi-open systems with a baseline migration rate (scenario III in Fig. 1B). For illustration purposes, we show the pre-IRS case, an example IRS under which the system approaches the transition regime, and another example IRS under which the system falls into the transition or slow-rebound regime. (A) As the system approaches the transition regime, the difference between PTS distributions across age groups is significantly reduced and even disappears. In particular, the two distributions almost completely overlap at the lowest mode around 0. The x-axis range for inset figures is 0-0.1. (B) MOI distribution starts to center around 1 with the majority of infections being either mono-clonal or multi-genomic with two genetically distinct parasites. These two molecular indicators are robust under implemented sampling schemes (sampling frequency and depth) and sampling limitations representative of those encountered in the collection of field data (the missing data issue and measurement error).

**Fig. S12.**
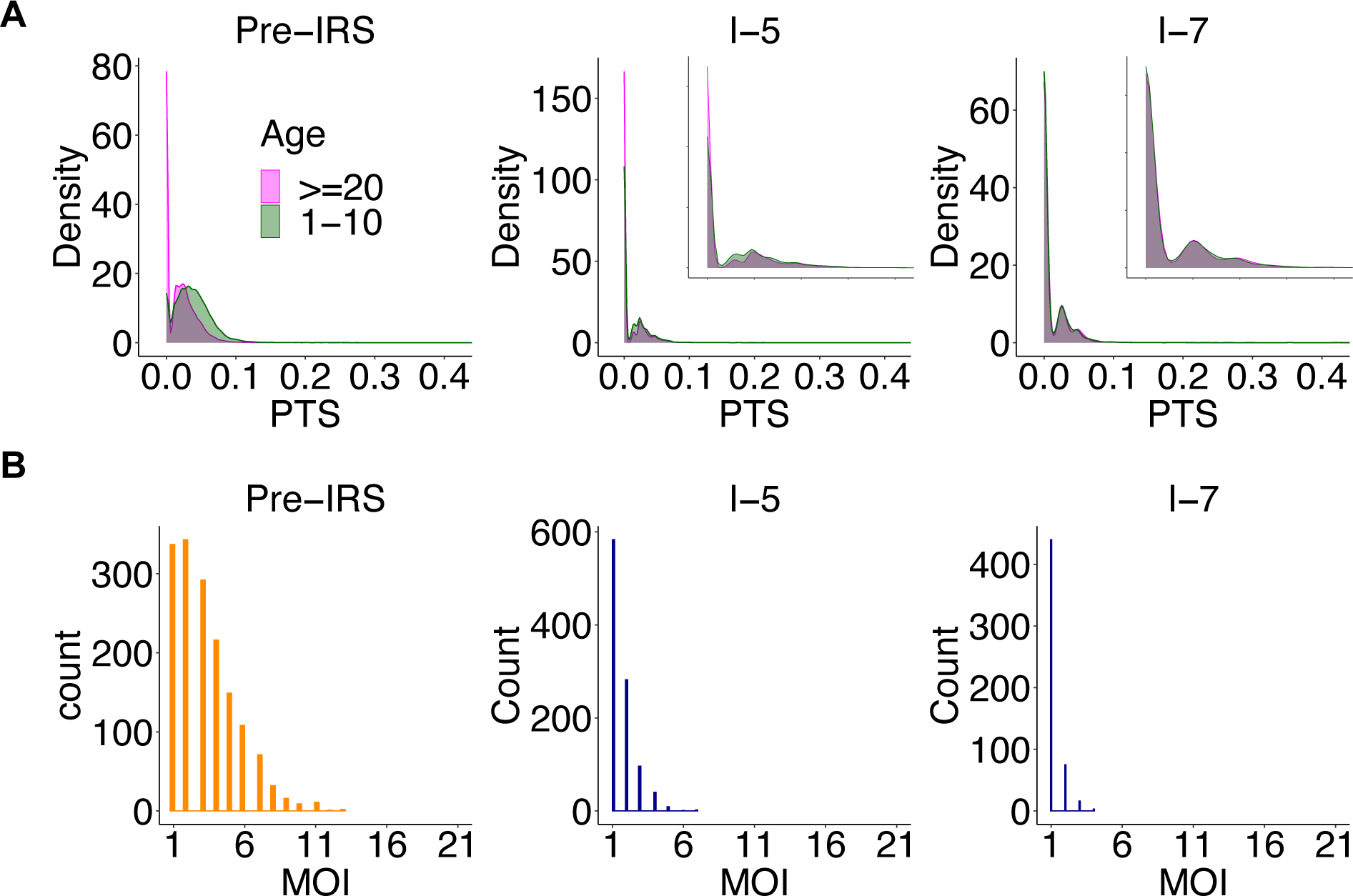
The two molecular indicators for non-seasonal semi-open systems with a baseline migration rate (scenario V in Fig. 1B). For illustration purposes, we show the pre-IRS case, an example IRS under which the system approaches the transition regime, and another example IRS under which the system falls into the transition or slow-rebound regime. (A) As the system approaches the transition regime, the difference between PTS distributions across age groups is significantly reduced and even disappears. In particular, the two distributions almost completely overlap at the lowest mode around 0. The x-axis range for inset figures is 0-0.1. (B) MOI distribution starts to center around 1 with the majority of infections being either mono-clonal or multi-genomic with two genetically distinct parasite. These two molecular indicators are robust under implemented sampling schemes (sampling frequency and depth) and sampling limitations representative of those encountered in the collection of field data (the missing data issue and measurement error).

**Fig. S13.**
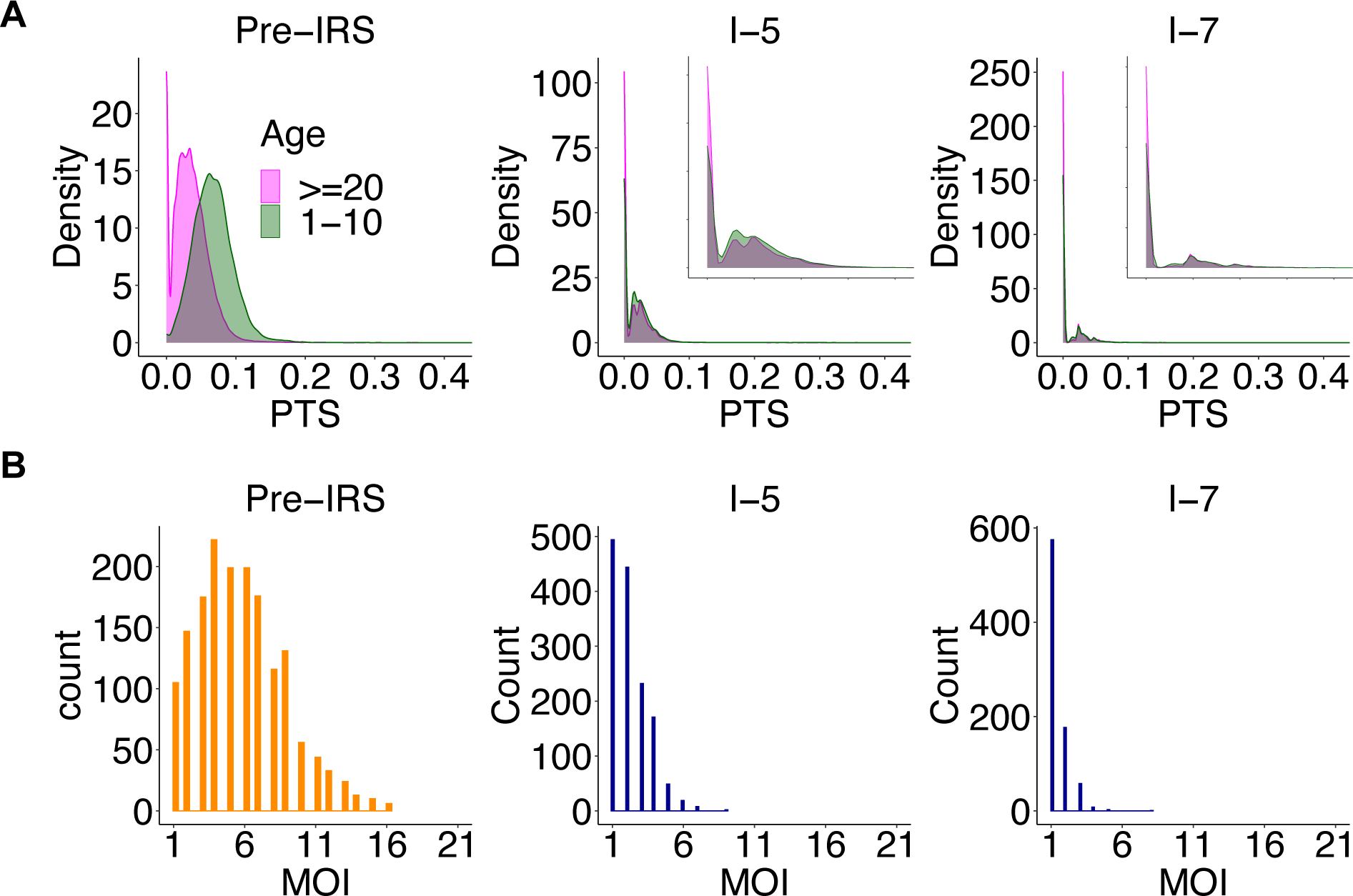
The two molecular indicators for seasonal semi-open systems with a high migration rate (scenario IV in Fig. 1B). For illustration purposes, we show the pre-IRS case, an example IRS under which the system approaches the transition regime, and another example IRS under which the system falls into the transition or slow-rebound regime. (A) As the system approaches the transition regime, the difference between PTS distributions across age groups is significantly reduced and even disappears. In particular, the two distributions almost completely overlap at the lowest mode around 0. The x-axis range for inset figures is 0-0.1. (B) MOI distribution starts to center around 1 with the majority of infections being either mono-clonal or multi-genomic with two genetically distinct parasites. These two molecular indicators are robust under implemented sampling schemes (sampling frequency and depth) and sampling limitations representative of those encountered in the collection of field data (the missing data issue and measurement error).

**Fig. S14.**
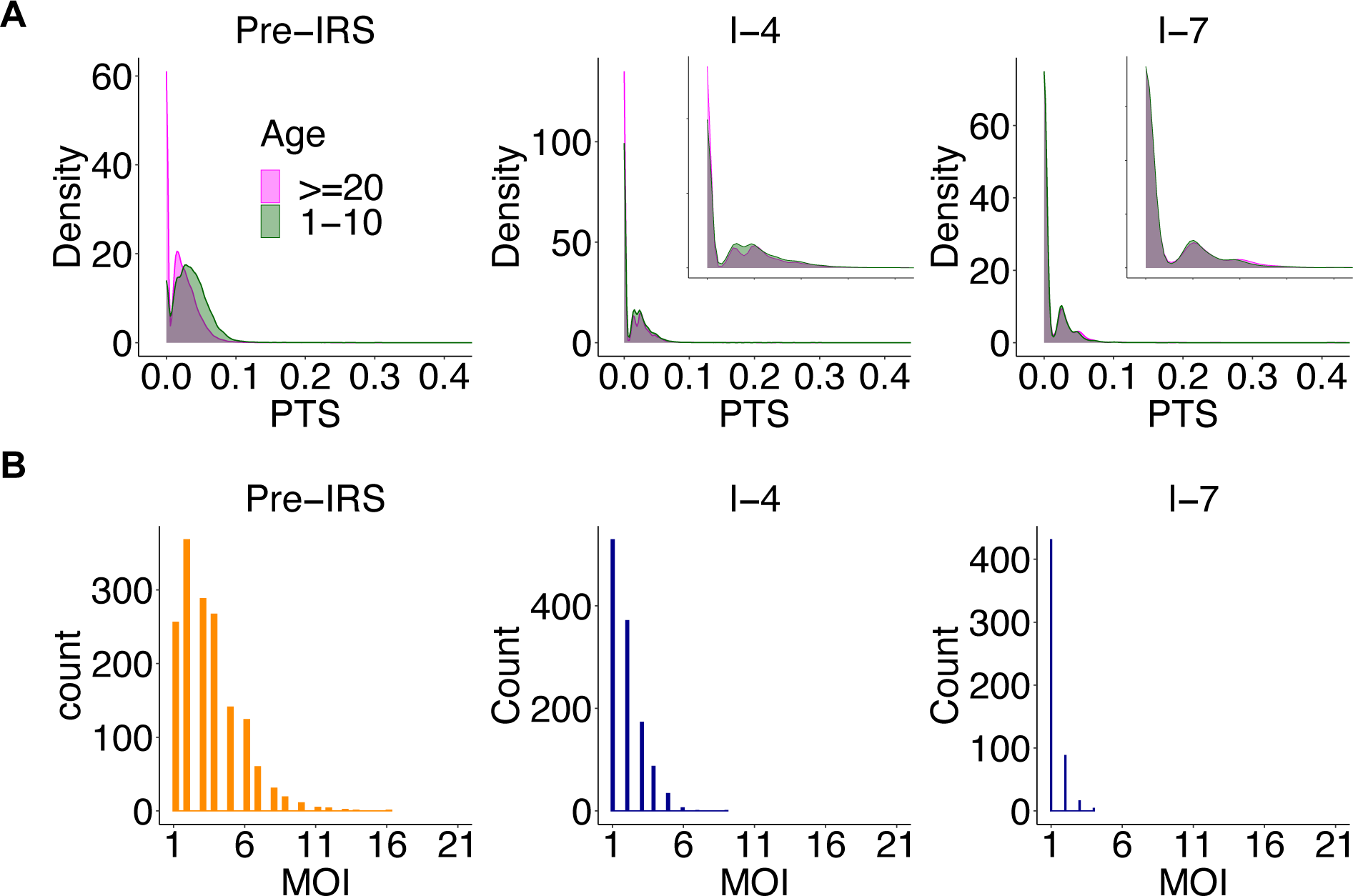
The two molecular indicators for non-seasonal semi-open system with a high migration rate (scenario VI in Fig. 1B). For illustration purposes, we show the pre-IRS case, an example IRS under which the system approaches the transition regime, and another example IRS under which the system falls into the transition or slow-rebound regime. (A) As the system approaches the transition regime, the difference between PTS distributions across age groups is significantly reduced and even disappears. In particular, the two distributions almost completely overlap at the lowest mode around 0. The x-axis range for inset figures is 0-0.1. (B) MOI distribution starts to center around 1 with the majority of infections being either mono-clonal or multi-genomic with two genetically distinct parasites. These two molecular indicators are robust under implemented sampling schemes (sampling frequency and depth) and sampling limitations representative of those encountered in the collection of field data (the missing data issue and measurement error).

**Fig. S15.**
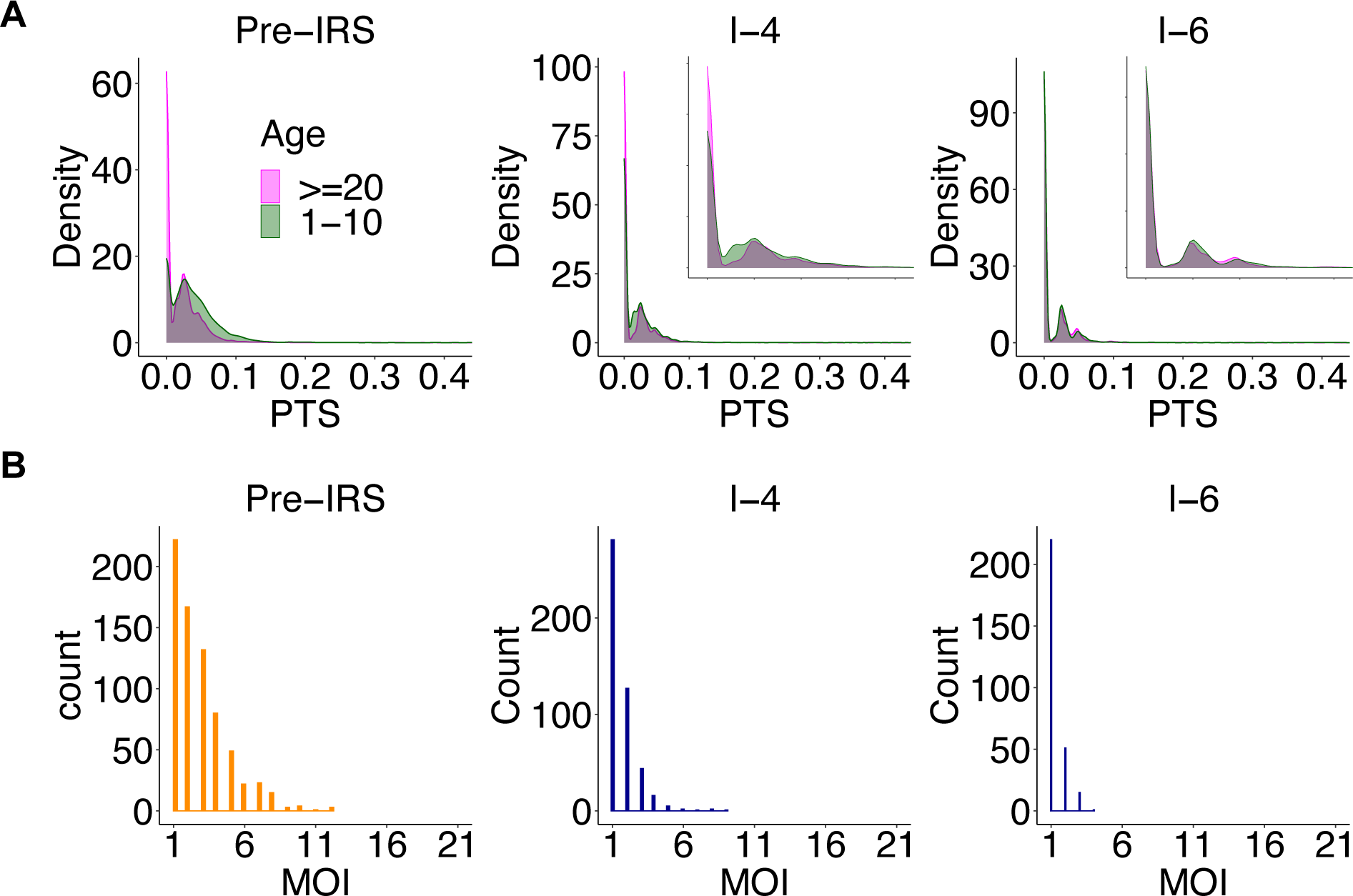
The two molecular indicators for non-seasonal regionally-open systems with a baseline migration rate and a medium regional pool (scenario XI in Fig. 1B). For illustration purposes, we show the pre-IRS case, an example IRS under which the system approaches the transition regime, and another example IRS under which the system falls into the transition or slow-rebound regime. (A) As the system approaches the transition regime, the difference between PTS distributions across age groups is significantly reduced and even disappears. In particular, the two distributions almost completely overlap at the lowest mode around 0. The x-axis range for inset figures is 0-0.1. (B) MOI distribution starts to center around 1 with the majority of infections being either mono-clonal or multi-genomic with two genetically distinct parasites. These two molecular indicators are robust under implemented sampling schemes (sampling frequency and depth) and sampling limitations representative of those encountered in the collection of field data (the missing data issue and measurement error).

**Fig. S16.**
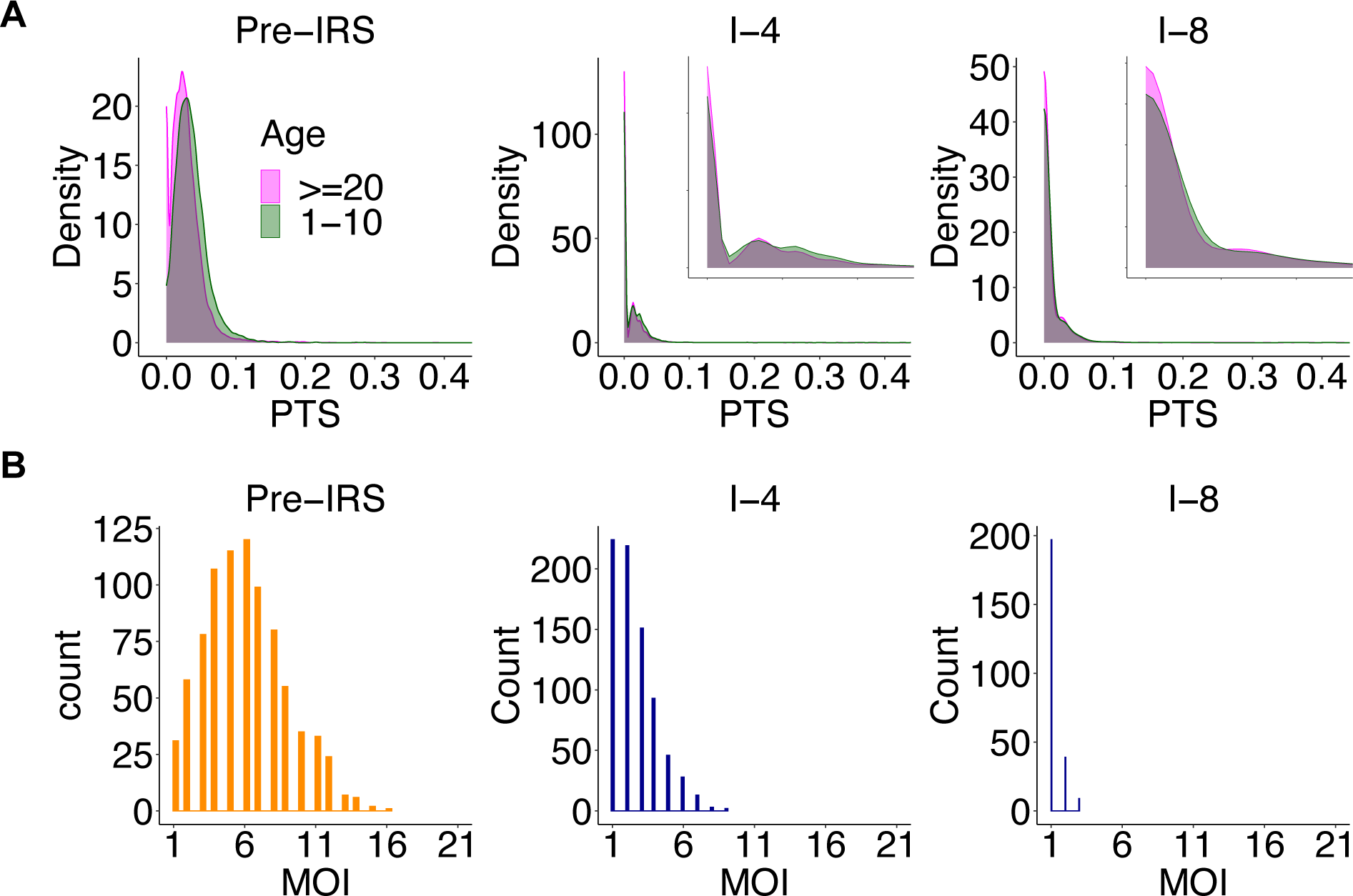
The two molecular indicators for seasonal regionally-open systems with a baseline migration rate and a large regional pool (scenario VIII in Fig. 1B). For illustration purposes, we show the pre-IRS case, an example IRS under which the system approaches the transition regime, and another example IRS under which the system falls into the transition or slow-rebound regime. (A) As the system approaches the transition regime, the difference between PTS distributions across age groups is significantly reduced and even disappears. In particular, the two distributions almost completely overlap at the lowest mode around 0. The x-axis range for inset figures is 0-0.05. (B) MOI distribution starts to center around 1 with the majority of infections being either mono-clonal or multi-genomic with two genetically distinct parasites. These two molecular indicators are robust under implemented sampling schemes (sampling frequency and depth) and sampling limitations representative of those encountered in the collection of field data (the missing data issue and measurement error).

**Fig. S17.**
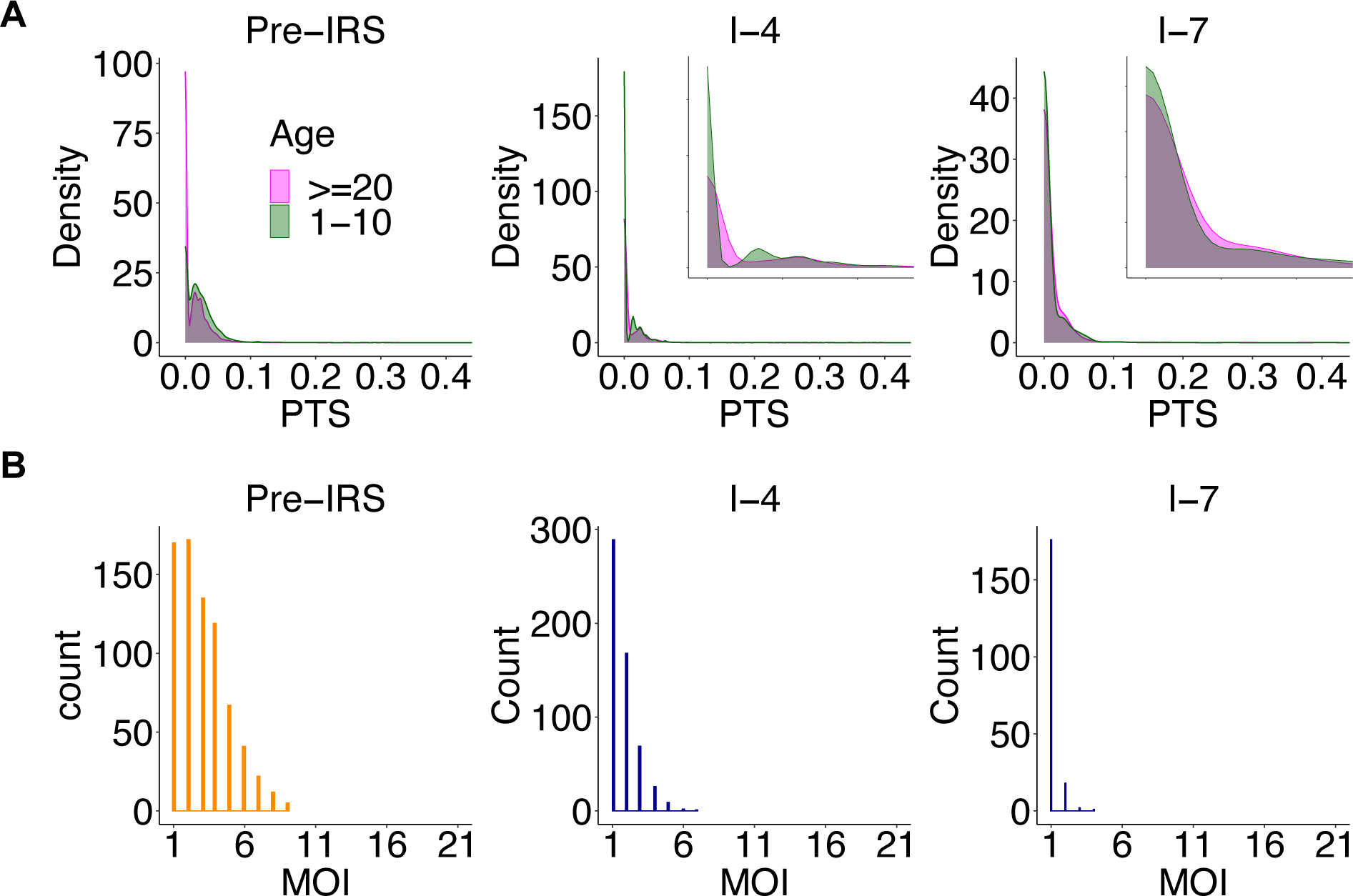
The two molecular indicators for non-seasonal regionally-open systems with a baseline migration rate and a large regional pool (scenario XII in Fig. 1B). For illustration purposes, we show the pre-IRS case, an example IRS under which the system approaches the transition regime, and another example IRS under which the system falls into the transition or slow-rebound regime. (A) As the system approaches the transition regime, the difference between PTS distributions across age groups is reversed. In particular, the PTS distribution of children develops an even higher peak around 0 than that of adults, the opposite of pre-IRS. The x-axis range for inset figures is 0-0.05. (B) MOI distribution starts to center around 1 with the majority of infections being either mono-clonal or multi-genomic with two genetically distinct parasites. These two molecular indicators are robust under implemented sampling schemes (sampling frequency and depth) and sampling limitations representative of those encountered in the collection of field data (the missing data issue and measurement error).

**Fig. S18.**
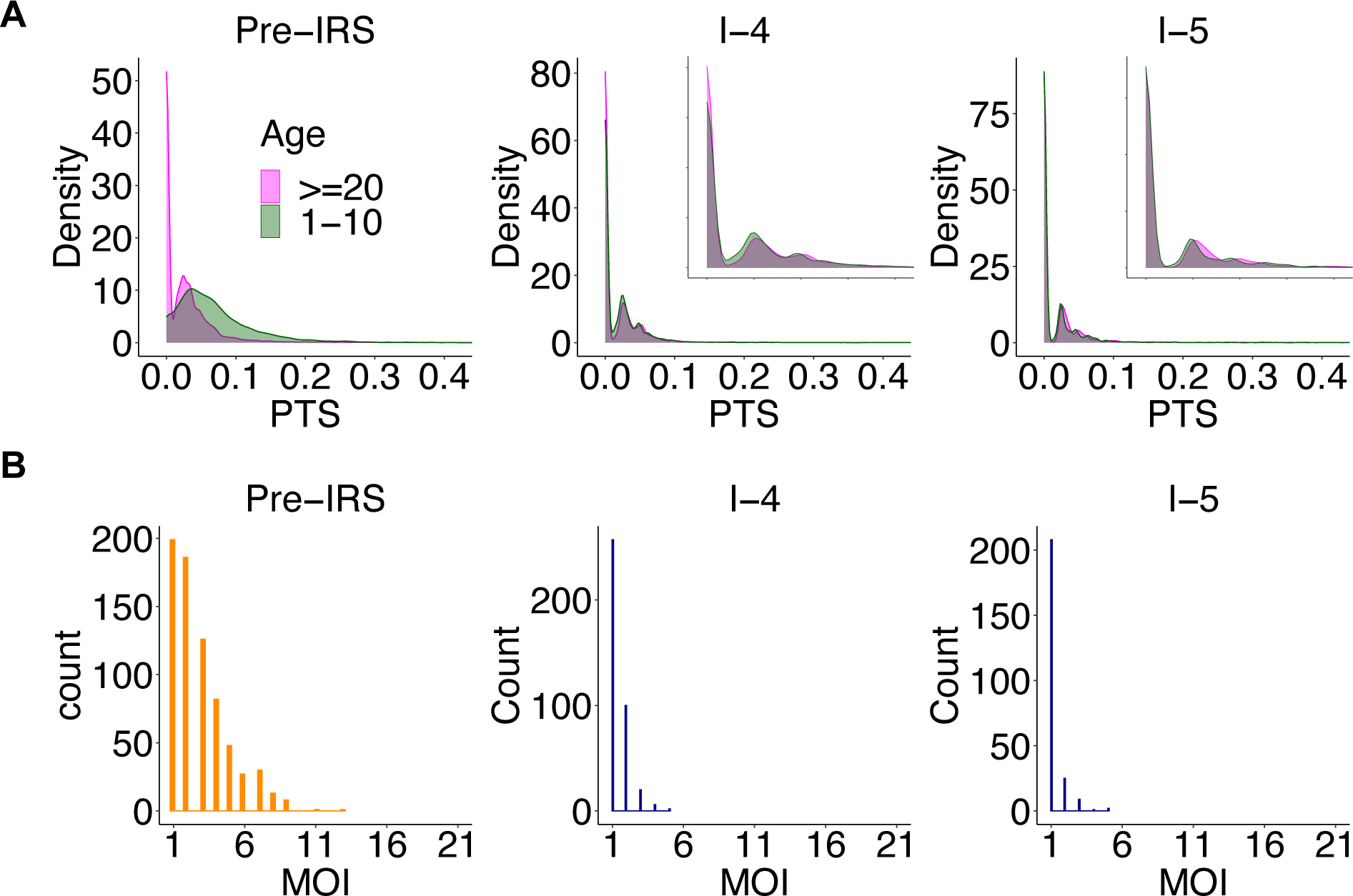
The two molecular indicators for seasonal regionally-open systems with a high migration rate and a medium regional pool (scenario IX in Fig. 1B). For illustration purposes, we show the pre-IRS case, an example IRS under which the system approaches the transition regime, and another example IRS under which the system falls into the transition or slow-rebound regime. (A) As the system approaches the transition regime, the difference between PTS distributions across age groups is significantly reduced and even disappears. In particular, the two distributions almost completely overlap at the lowest mode around 0. The x-axis range for inset figures is 0-0.1. (B) MOI distribution starts to center around 1 with the majority of infections being either mono-clonal or multi-genomic with two genetically distinct parasites. These two molecular indicators are robust under implemented sampling schemes (sampling frequency and depth) and sampling limitations representative of those encountered in the collection of field data (the missing data issue and measurement error).

**Fig. S19.**
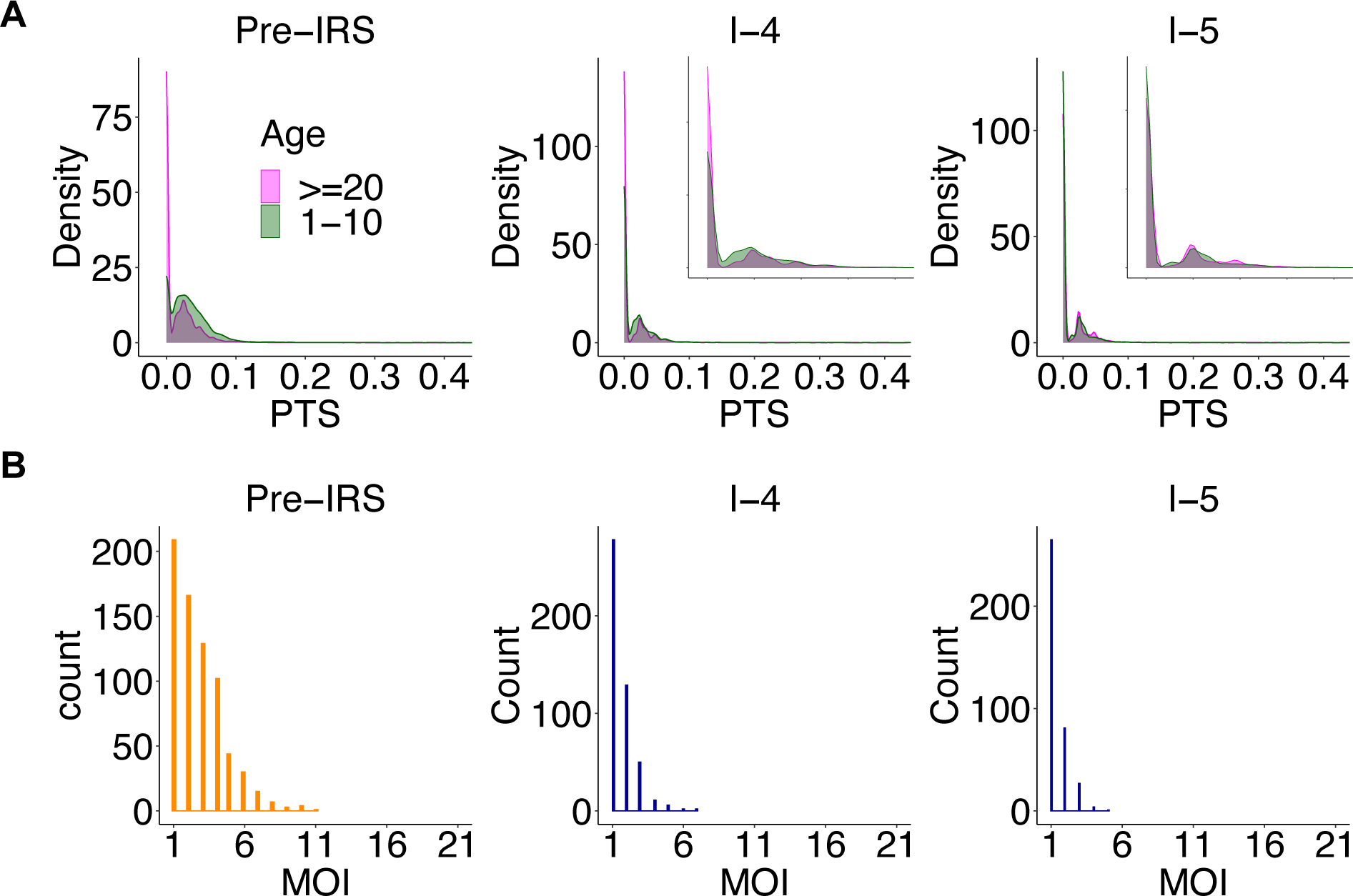
The two molecular indicators for non-seasonal regionally-open systems with a high migration rate and a medium regional pool (scenario XIII in Fig. 1B). For illustration purposes, we show the pre-IRS case, an example IRS under which the system approaches the transition regime, and another example IRS under which the system falls into the transition or slow-rebound regime. (A) As the system approaches the transition regime, the difference between PTS distributions across age groups is significantly reduced and even disappears. In particular, the two distributions almost completely overlap at the lowest mode around 0. The x-axis range for inset figures is 0-0.1. (B) MOI distribution starts to center around 1 with the majority of infections being either mono-clonal or multi-genomic with two genetically distinct parasites. These two molecular indicators are robust under implemented sampling schemes (sampling frequency and depth) and sampling limitations representative of those encountered in the collection of field data (the missing data issue and measurement error).

**Fig. S20.**
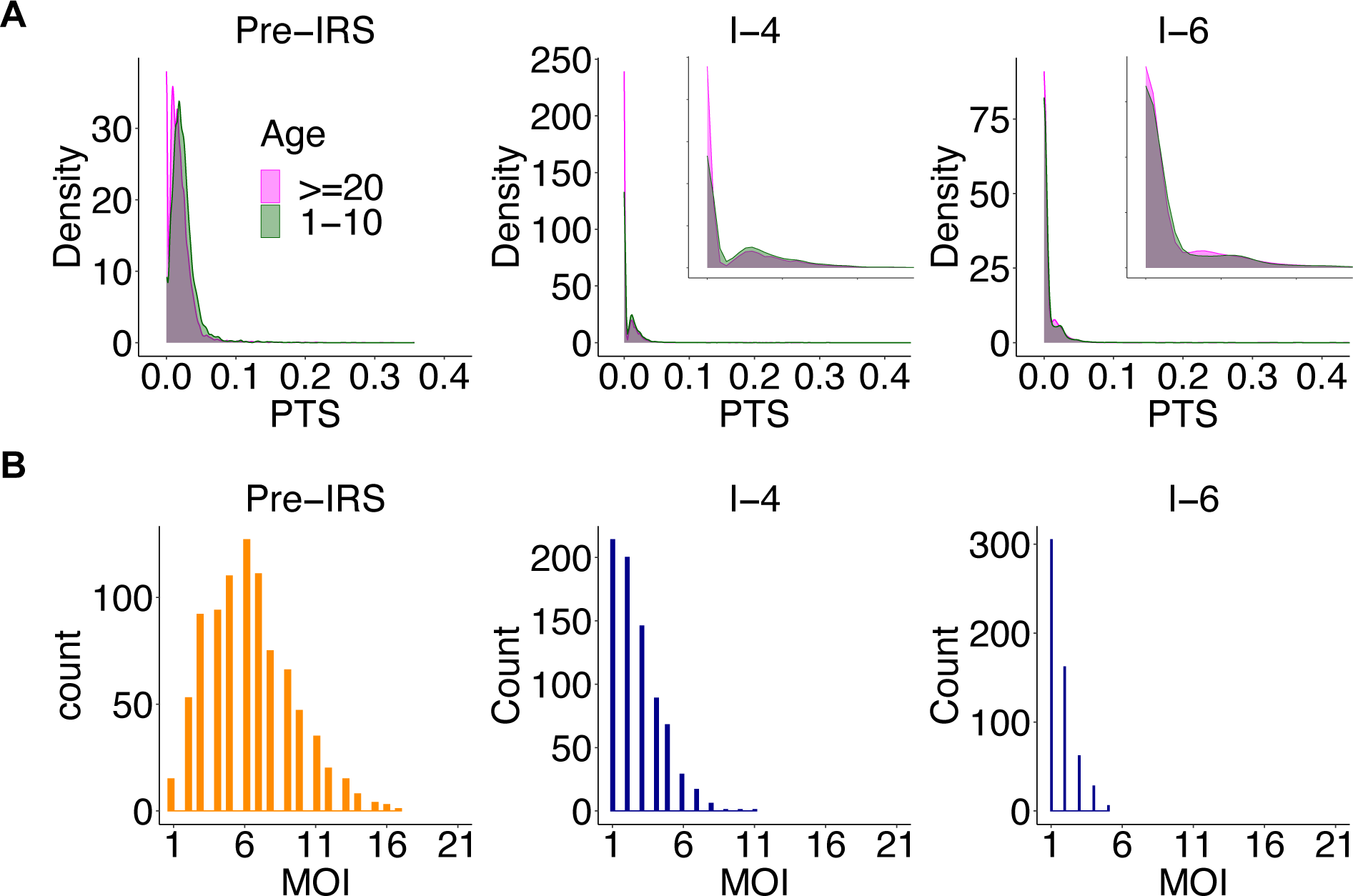
The two molecular indicators for seasonal regionally-open systems with a high migration rate and a large regional pool (scenario X in Fig. 1B). For illustration purposes, we show the pre-IRS case, an example IRS under which the system approaches the transition regime, and another example IRS under which the system falls into the transition or slow-rebound regime. (A) As the system approaches the transition regime, the difference between PTS distributions across age groups is significantly reduced and even disappears. In particular, the two distributions almost completely overlap at the lowest mode around 0. The x-axis range for inset figures is 0-0.05. (B) MOI distribution starts to center around 1 with the majority of infections being either mono-clonal or multi-genomic with two genetically distinct parasites. These two molecular indicators are robust under implemented sampling schemes (sampling frequency and depth) and sampling limitations representative of those encountered in the collection of field data (the missing data issue and measurement error).

**Fig. S21.**
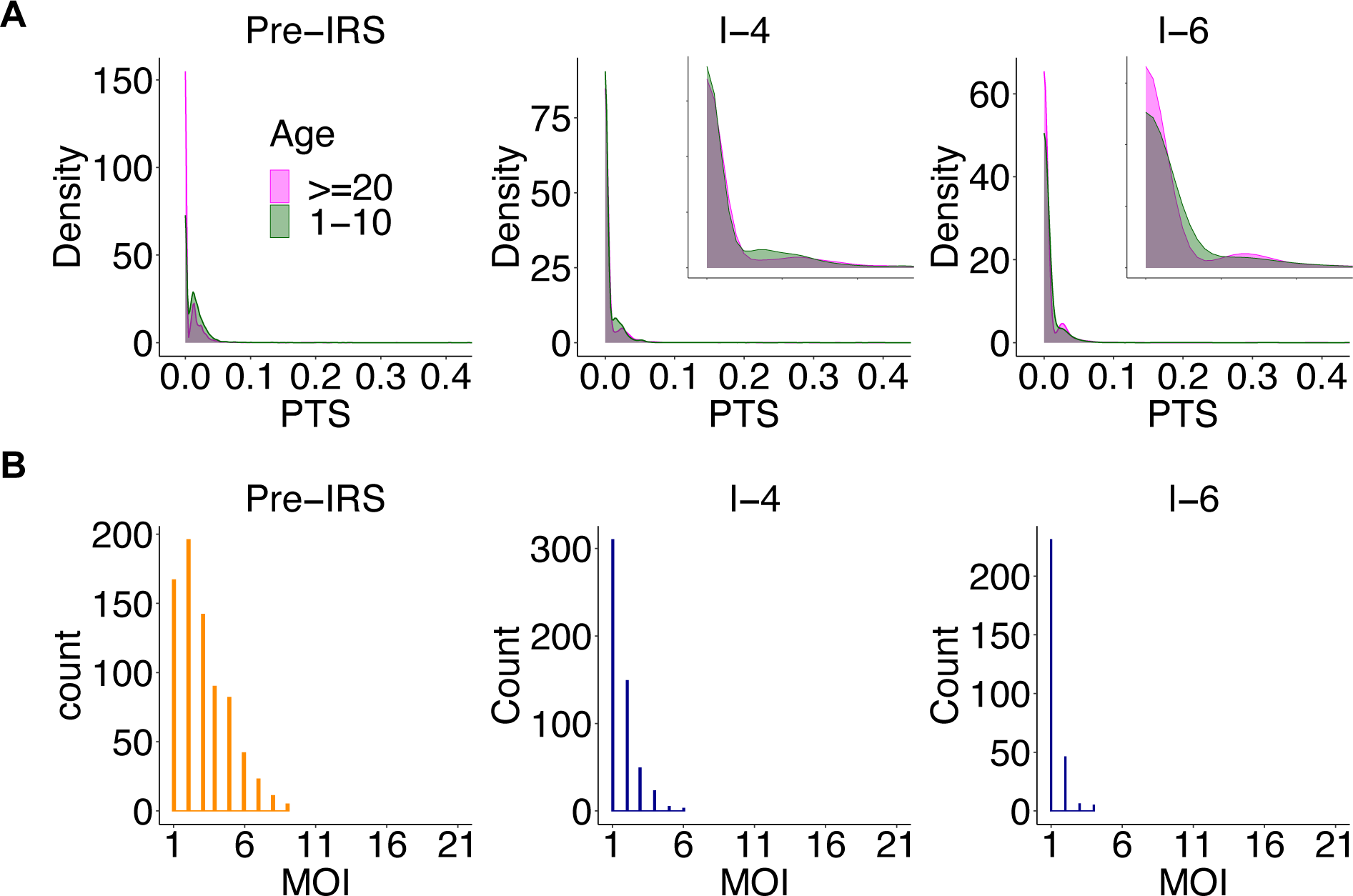
The two molecular indicators for non-seasonal regionally-open systems with a high migration rate and a large regional pool (scenario XIV in Fig. 1B). For illustration purposes, we show the pre-IRS case, an example IRS under which the system approaches the transition regime, and another example IRS under which the system falls into the transition or slow-rebound regime. (A) As the system approaches the transition regime, the difference between PTS distributions across age groups is significantly reduced and even disappears. In particular, the two distributions almost completely overlap at the lowest mode around 0. The x-axis range for inset figures is 0-0.05. (B) MOI distribution starts to center around 1 with the majority of infections being either mono-clonal or multi-genomic with two genetically distinct parasites. These two molecular indicators are robust under implemented sampling schemes (sampling frequency and depth) and sampling limitations representative of those encountered in the collection of field data (the missing data issue and measurement error).

**Fig. S22.**
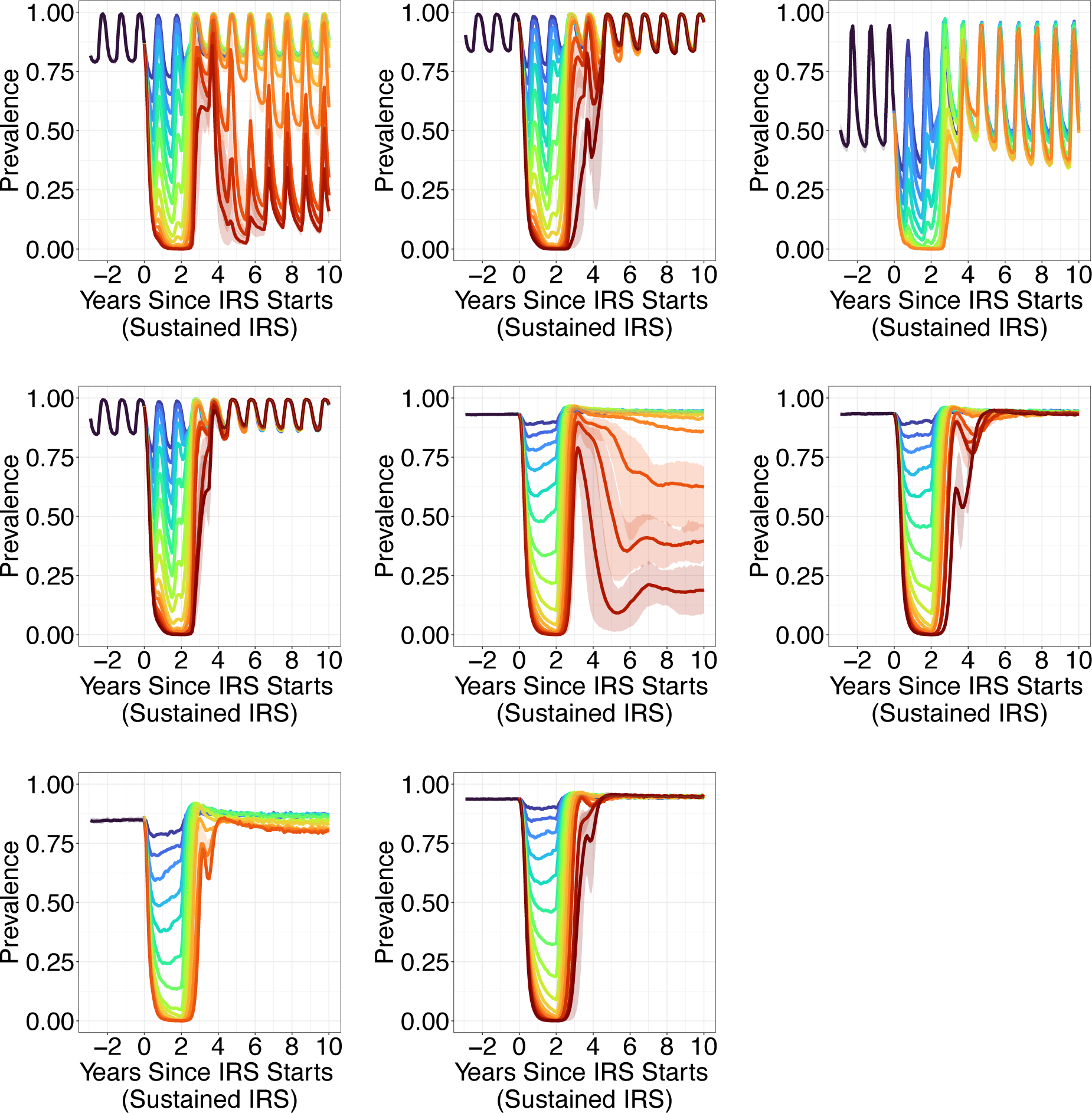
Rebound dynamics of prevalence across time, under a series of transient IRS interventions (2 years). The dynamics for closed and semi-open systems can be categorized into the three regimes of fast-rebound, transition, and slow-rebound regimes. However, regionally-open systems always rebound rapidly. The top row from left to right are seasonal and semi-open systems with a high migration rate (scenario IV in Fig. 1B), seasonal and regionally-open systems with a baseline migration rate and a large pool (scenario VIII), seasonal and regionally-open systems with a high migration rate and a medium size pool (scenario IX). The middle row from left to right are seasonal and regionally-open systems with a high migration rate and a large pool (scenario X), non-seasonal and semi-open systems with a high migration rate (scenario VI), non-seasonal and regionally-open systems with a baseline migration rate and a large pool (scenario XII). The bottom row are non-seasonal and regionally-open systems with a high migration rate and a medium size pool (scenario XIII), non-seasonal and regionally-open systems with a high migration rate and a large pool (scenario XIV). Simulation runs with other parameter values are shown in Fig. 6A.

